# GP73 is a glucogenic hormone regulating SARS-CoV-2-induced hyperglycemia

**DOI:** 10.1101/2021.04.30.21256060

**Authors:** Luming Wan, Huan Yang, Huilong Li, Jing Gong, Yongqiang Deng, Yuehua Ke, Xiaopan Yang, Haotian Lin, Yanhong Zhang, Changjun Wang, Dongyu Li, Huapeng Wang, Yumeng Peng, Qiulin Yan, Linfei Huang, Xiaolin Wang, Qi Gao, Changqing Lin, Fei Zheng, Lei Xu, Jialong Liu, Xuemiao Zhang, Xiaoli Yang, Chengfeng Qin, Zhiwei Sun, Feixiang Wu, Congwen Wei, Hui Zhong

**Affiliations:** Beijing Institute of Biotechnology, Beijing, 100850, China; Guangxi liver cancer Diagnosis and Treatment Engineering and Technology research center, Nanning, China; Beijing Institute of Microbiology and Epidemiology, Beijing, 100850, China; Center for Disease Control and Prevention of PLA, Beijing, China; Hotgen Biotech Co., Ltd. Beijing, China; Department of Clinical Laboratory, the Third Medical Centre, Chinese PLA General Hospital, China

**Author notes:** These authors contributed equally to this work (Luming Wan, Huan Yang, Huilong Li, Jing Gong, Yongqiang Deng, Yuehua Ke). These authors jointly supervised the work: Xiaoli Yang, Chengfeng Qin, Zhiwei Sun, Feixiang Wu, Congwen Wei, and Hui Zhong.

**Keywords:** SARS-COV-2, hepatic gluconeogenesis, GP73, hyperglycemia, diabetes

## Abstract

Severe acute respiratory syndrome coronavirus 2 (SARS-CoV-2) infection induces new-onset diabetes and severe metabolic complications of pre-existing diabetes. The pathogenic mechanism underlying this is incompletely understood. Here, we provided evidence linking circulating GP73 with the exaggerated gluconeogenesis triggered by SARS-CoV-2 infection. We found that SARS-CoV-2 infection or glucotoxic condition increased the cellular secretion of GP73. Secreted GP73 trafficked to the liver and kidney to stimulate gluconeogenesis through cAMP/PKA pathway. By using global phosphoproteomics, we found a drastic remodeling of PKA kinase hub exerted by GP73. Notably, COVID-19 patients showed pathologically elevated plasma GP73, and neutralization of the secreted GP73 inhibited enhanced PKA signaling and glucose production associated with SARS-CoV-2 infection. GP73 blockade also reduced gluconeogenesis and lowered hyperglycemia in type 2 (T2D) diabetic mice. Therefore, our findings provide novel insight into the roles of GP73 as a key glucogenic hormone and mechanistic clues underlying the development of SARS-CoV-induced glucose abnormalities.

## Introduction

Under physiological circumstances, blood glucose levels are maintained within a narrow range to protect the body against hypoglycemia during fasting and excessive hyperglycemia following a high-carbohydrate meal(Roder et al., 2016). Glucose homeostasis is achieved primarily via hormonal modulation of glucose production by the liver(Sharabi et al., 2015) and glucose uptake by skeletal muscle, heart muscle, and fat(Chadt and Al-Hasani, 2020; Kowalski and Bruce, 2014). Hepatic glucose production (HGP) involves a combination of glycogen breakdown (glycogenolysis) and de novo synthesis of glucose from noncarbohydrate precursors (gluconeogenesis)(Sharabi et al., 2015). Glycogenolysis occurs rapidly in the early stage of the fed-to-fasting transition, and gluconeogenesis is the main contributor to hepatic and renal glucose production during prolonged fasting states(Chung et al., 2015). The rate of gluconeogenic flux is controlled by the activities of key unidirectional enzymes, such as pyruvate carboxylase (PCX), phosphoenolpyruvate carboxykinase 1 (PCK1), fructose 1,6-bisphosphatase (FBP1), and glucose-6-phosphatase (G6Pase)(Zhang et al., 2018). The genes encoding these proteins are strongly controlled at the transcriptional level by key hormones including insulin, glucagon, and glucocorticoids(Fleig et al., 1984). These protein hormones are generally produced via the cleavage of a larger proprotein in response to nutrient-derived cues or glucose and trafficked to the target organ via the circulation upon secretion. These hormones stimulate rapid signal transduction via a second messenger system through a cell-surface receptor on target cells(Zhang et al., 2018). Excessive HGP not only contributes to exaggerated fasting and postprandial hyperglycemia in type 1 diabetes (T1D) and type 2 diabetes (T2D) patients, but also contributes to stress-, infection-, and inflammation-associated hyperglycemia(Deng et al., 2011; Petersen et al., 2017).

Severe acute respiratory syndrome coronavirus 2 (SARS-CoV-2), which caused the coronavirus disease 2019 (COVID-19) pandemic, primarily invades pulmonary epithelial cells via the surface angiotensin converting enzyme 2 (ACE2) receptor and eventually leads to respiratory symptoms(Hoffmann et al., 2020). The clinical presentation of COVID-19 ranges from an asymptomatic or mild disease to severe respiratory symptoms and death. Pre-existing diabetes, atherosclerosis, and other health conditions substantially increase COVID-19 mortality and morbidity (Ceriello et al., 2020; Chen et al., 2020). Of note, diabetic is not only a risk factor for the severe form of COVID-19 disease, but also that infection can induce new-onset diabetes and severe metabolic complications in patients with preexisting diabetes(Cristelo et al., 2020; Lim et al., 2021; Sathish and Chandrika Anton, 2021). Exceptionally high doses of insulin are required to achieve normal blood glucose levels (Jornayvaz et al., 2020). Although impaired insulin secretion due to pancreatic damage plays an important role in the development of COVID-19(Alves et al., 2020; Liu et al., 2020), the pathogenic mechanisms underlying new-onset T2D are largely unknown.

GP73 is a type II transmembrane Golgi protein located at the luminal surface of the Golgi apparatus, consisting of a short N-terminal cytoplasmic domain, a transmembrane domain, and a large C-terminal domain. GP73 cycles out of the cis-Golgi to endosomes for cleavage by the convertase furin, which results in its release into the extracellular space(Hu et al., 2011; Puri et al., 2002). Circulating GP73 is implicated in the regulation of cell-to-cell communication triggered by the unfolded protein response(Wei et al., 2019). The present study provides evidence that GP73 is a fasting-induced hormone that promotes hepatic and renal glucose production in endocrine and autocrine manners. Notably, SARS-CoV-2 infection and hyperglycemia greatly increased the secretion of GP73. Therefore, targeting GP73 might be a potential therapeutic strategy for patients with SARS-CoV-2 infection and diabetes who present alterations in the levels of this hormone.

## Results

### GP73 secretion is promoted during SARS-CoV-2 infection

The fact that hepatocyte GP73 expression is upregulated in patients with viral and nonviral liver disease suggests that this protein is triggered during hepatocyte injury (Gatselis et al., 2020). The liver damage caused by SARS-CoV-2 infection prompted us to investigate whether infection with this virus altered GP73 expression(Alqahtani and Schattenberg, 2020). Notably, SARS-CoV-2-infected patients had approximately 2-fold higher circulating GP73 levels than the reference population (P <0.0001; Table 1 and Figure 1A). When disease status was classified into mild, moderate, and severe, the serum GP73 concentration was significantly higher in patients with severe or moderate disease than patients with moderate or mild disease (Table 2 and Figure 1B).

**Table 1.** Clinical characteristics of COVID-19 patients and reference controls.

| Parameter | COVID-19<br>(n=136) | Reference<br>(n=100) | P value |
| --- | --- | --- | --- |
| **Clinical characteristics** |  |  |  |
| Age (y) | 60 ± 14 | 58 ± 15 | 0.2939 |
| Male (n) | 78 | 59 |  |
| Female (n) | 58 | 41 |  |
| **Laboratory examination** |  |  |  |
| Leukocytes (10<sup>9</sup>/L) | 6.47 ± 5.15 | 5.13 ± 2.29 | 0.0158 |
| Lymphocytes (10<sup>9</sup>/L) | 1.56 ± 0.67 | 1.31 ± 0.45 | 0.0014 |
| Neutrophils (10<sup>9</sup>/L) | 4.09 ± 1.99 | 3.05 ± 1.56 | < 0.0001 |
| Monocytes (10<sup>9</sup>/L) | 0.45 ± 0.15 | 0.41 ± 0.21 | 0.0891 |
| Platelets (10<sup>9</sup>/L) | 209.44 ± 66.69 | 215.56 ± 76.67 | 0.5140 |
| CRP (mg/L) | 22.91 ± 11.98 | 5.41 ± 2.65 | < 0.0001 |
| ALT (40 U/L) | 34.96 ± 29.09 | 18.61 ± 9.58 | < 0.0001 |
| AST (40 U/L) | 26.88 ± 18.27 | 16.16 ± 4.80 | < 0.0001 |
| BG (mmol/L) | 102.27 ± 32.97 | 88.21 ± 7.73 | 0.0409 |
| Abbreviations: CRP, C-reactive protein; ALT, alanine transaminase; AST, Aspartate transaminase; BG, blood glucose.<br>P value was calculated using the Mann-Whitney U test for continuous variables between two groups. |  |  |  |

**Fig. 1.**
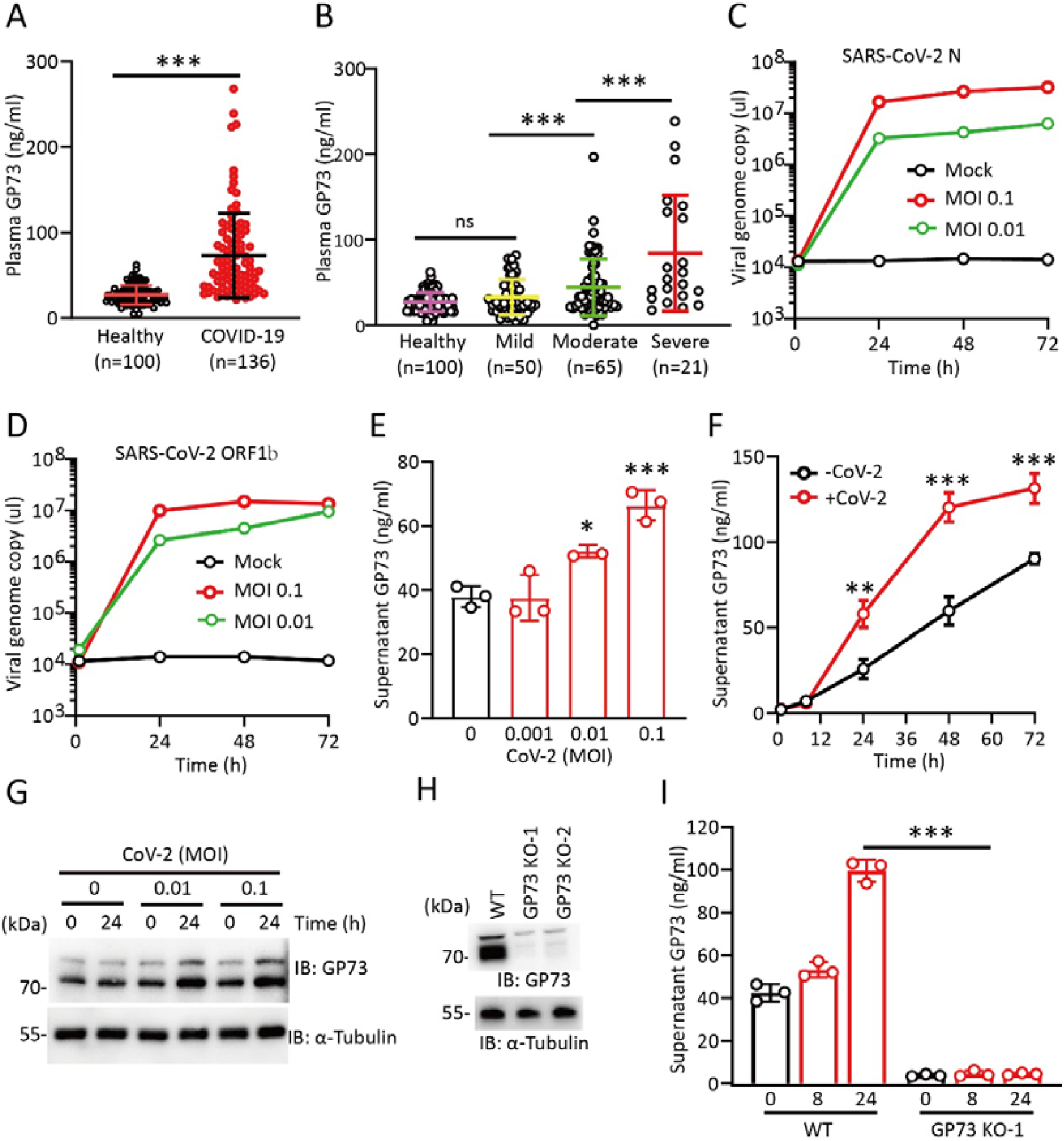
GP73 secretion is promoted during SARS-CoV-2 infection. (A) Plasma GP73 levels in healthy controls (n=100) and COVID-19 patients (n=136).***P < 0.001; Mann-Whitney U test. (B) Plasma GP73 levels in COVID-19 patients with different disease severities. ***, P < 0.001. (C-D) QRT-PCR analysis of SARS-CoV-2 genome copy number per μL using specific primers for N (C) or ORF1b (D) in the supernatants of Huh-7 cells infected with SARS-CoV-2 for the indicated times and MOI. (E-F) ELISA analysis of GP73 levels in the supernatants of Huh-7 cells infected with SARS-CoV-2 (CoV-2) at the indicated MOIs for 24 h (E) or with SARS-CoV-2 (MOI 0.1) for the indicated times (F). (G) Immunoblotting analysis of intracellular GP73 levels in Huh-7 cells infected with SARS-CoV-2 (CoV-2) at the indicated MOIs for 24 h. (H) Immunoblotting analysis of intracellular GP73 levels in WT or GP73 KO-1 Huh-7 cells. (I) ELISA analysis of GP73 levels in the supernatants of WT or GP73 KO-1 Huh-7 cells infected with SARS-CoV-2 (MOI 0.1) for the indicated times. Cell-based studies were performed independently at least three times with comparable results. The data are presented as the means ± SEMs. *, P < 0.05; **, P < 0.01; and ***, P < 0.001; one-way or two-way ANOVA followed by Bonferroni’s post hoc test for B-F, I.

**Table 2.** Clinical characteristics of COVID-19 patients with disease severity.

| **Parameter** | **Mild**<br>**(n=50)** | **Moderate**<br>**(n=65)** | **Severe**<br>**(n=21)** | **P value** |
| --- | --- | --- | --- | --- |
| **Clinical characteristics** |  |  |  |  |
| Age (y) | 54±13 | 61±12 | 65±10 | 0.0007 |
| Male (n) | 27 | 36 | 16 |  |
| Female (n) | 23 | 29 | 5 |  |
| **Laboratory examination** |  |  |  |  |
| Leukocytes (10<sup>9</sup>/L) | 4.55 ±2.09 | 6.77 ± 2.02 | 8.63 ± 3.08 | < 0.0001 |
| Lymphocytes (10<sup>9</sup>/L) | 1.75 ± 0.57 | 1.64 ± 0.65 | 1.00 ± 0.63 | < 0.0001 |
| Neutrophils (10<sup>9</sup>/L) | 3.32 ± 0.95 | 3.77 ± 1.49 | 6.37 ± 2.85 | < 0.0001 |
| Monocytes (10<sup>9</sup>/L) | 0.43 ± 0.13 | 0.47 ± 0.16 | 0.42 ± 0.18 | 0.2580 |
| Platelets (10<sup>9</sup>/L) | 215.3 ± 55.5 | 217.1 ± 59.5 | 176.4 ± 93.2 | 0.0361 |
| CRP (mg/L) | 7.08 ± 3.04 | 18.46 ± 9.79 | 38.9 ± 28.43 | < 0.0001 |
| ALT (40 U/L) | 28.08 ± 17.2 | 38.89 ± 34.1 | 34.5 ± 27.32 | 0.1315 |
| AST (40 U/L) | 22.00 ± 9.10 | 27.1 ± 16.67 | 34.2 ± 29.56 | 0.0224 |
| BG (mmol/L) | 90.0 ± 21.18 | 101.2 ± 29.8 | 134.5 ± 44.0 | <0.0001 |
| Abbreviations: CRP, C-reactive protein; ALT, alanine transaminase; AST, Aspartate transaminase; BG, blood glucose.<br>P value was calculated using Fisher's exact test or $\chi^2$ test for categorical variables. | | | | |

To assess whether cultured hepatocytes are capable of generating and secreting GP73, human hepatocyte Huh-7 cells were infected with SARS-CoV-2, and GP73 secretion was examined over time during infection. Successful infection was confirmed by the observation of increasing extracellular viral RNA levels over the course of the infection period (Figure 1C-D). Interestingly, cultured hepatocytes exposed to SARS-CoV-2 exhibited a robust time-and dose-dependent increase in GP73 secretion (Figure 1E-F). Intracellular GP73 protein was also increased in SARS-CoV-2-infected cells (Figure 1G). In particular, we did not observe any detectable GP73 accumulation following SARS-CoV-2 infection of Huh-7 cells lacking GP73 (GP73 KO-1; Figure 1H-I). Together, these results demonstrate that SARS-CoV-2 infection promotes GP73 production and secretion.

### Increase in circulating GP73 elevates fasting blood glucose

To understand the acute response to elevated circulating GP73 levels, we expressed and purified the mouse GP73 c-terminal fragment (GP73 52-401; rmGP73) with His-Tag (Figure S1A). We developed a mouse GP73 sandwich ELISA and constructed a standard curve using rmGP73 to calculate plasma GP73 levels in mice (Figure S1B). To evaluate the specificity of this ELISA kit, GP73 knockout (GP73 KO) mice were generated using CRISPR-Cas9 by excising exon 4 of the GP73 gene (NP105348). The GP73 transcript was not detected in multiple tissues from mutant mice. The mouse GP73 sandwich ELISA exhibited high specificity in plasma from WT and GP73 KO mice (Figure S1C). Administration of a single dose of 0.1 mg/kg rmGP73 led to an approximately 2-fold increase in the levels of plasma GP73 compared to control mice (Figure S1D). Plasma His-tagged rmGP73 had a half-life of approximately 30 min, and a peak level was achieved 15 min post-injection (Figure S1E). We then experimentally elevated plasma GP73 levels via the intravenous (i.v.) injection of 0.1 mg/kg rmGP73 into mice that were subjected to a 12-h fast, and plasma glucose levels were measured at 15, 30, 60, 90, and 120 min post-injection. In this setting, rmGP73 induced an immediate spike in blood glucose levels (Figure 2A). A GP73-specific antibody completely blocked the glucose-stimulating effect of recombinant GP73 (Figure 2B). It should be noted that compensatory hyperinsulinemia (measured at the 15-min time point, Figure 2C) was not sufficient to normalize blood glucose levels by 48 h post-injection, as mice receiving rmGP73 still displayed higher blood glucose levels 24 and 48 h after the injection (Figure 2D). We then injected rmGP73 daily for three days. Repeated GP73 injection under this condition produced a sharp rise in blood glucose levels (Figure 2E).

**Fig. 2.**
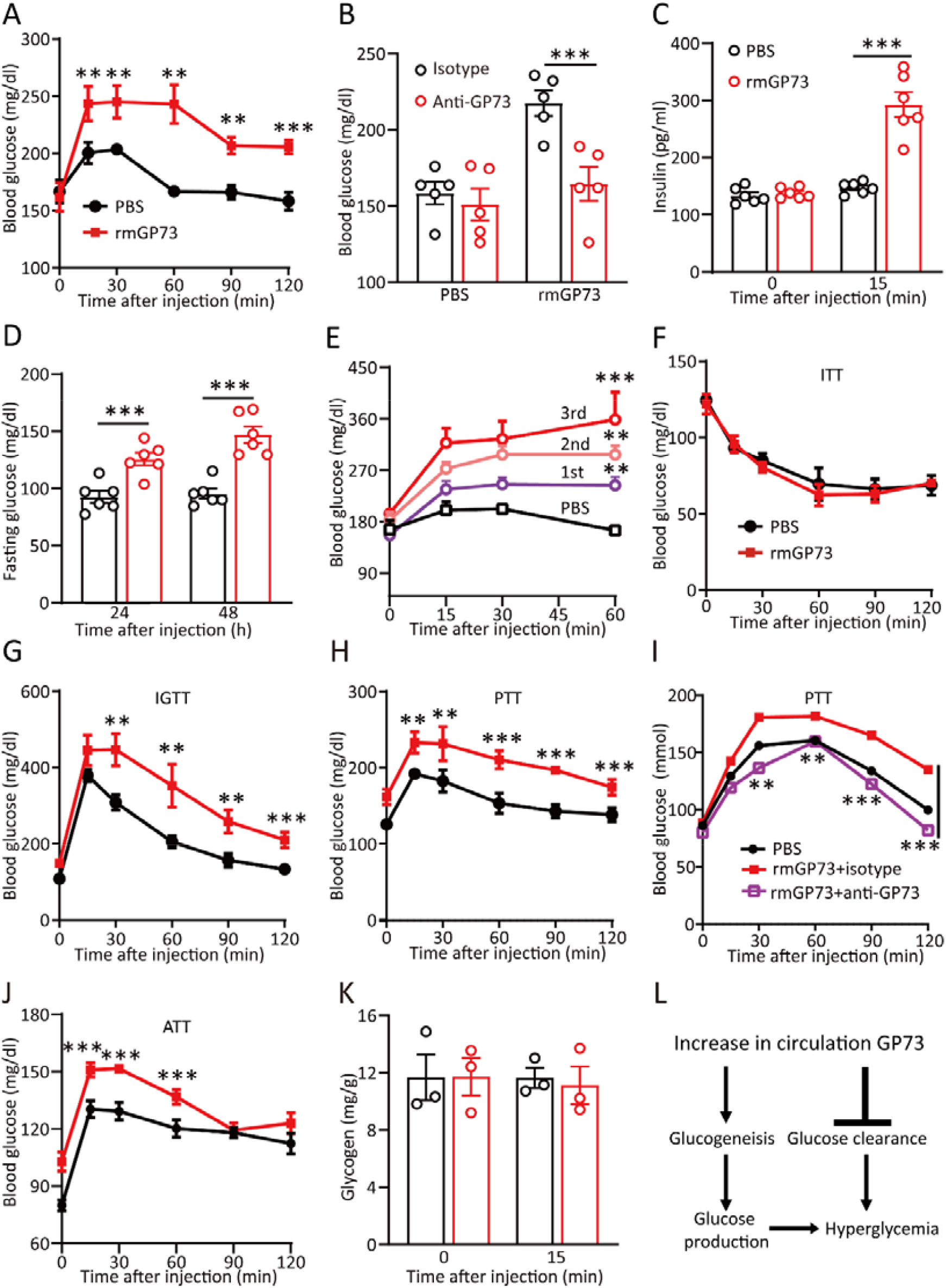
Increase in circulating GP73 elevates fasting blood glucose. (A) Plasma glucose levels were measured at the indicated times after a single dose of intravenous (i.v.) rmGP73 (0.1 mg/kg) or PBS in mice subjected to a 12 h fast prior to injection (n = 6 in each group). (B) Plasma glucose levels were measured 15 min after rmGP73 injection with IgG isotype or an anti-GP73 antibody (n = 5 in each group). (C) Plasma insulin levels were measured 15 min after rmGP73 injection (n = 6 in each group). (D) Plasma glucose levels were measured 24 h or 48 h after rmGP73 injection (n = 6 in each group). (E) Plasma glucose levels were measured at the indicated times after rmGP73 injection once daily for three days (n = 6 in each group). (F-J) Insulin tolerance test (ITT; F), glucose tolerance test (GTT; G), pyruvate tolerance test (PTT; H), PTT with or without anti-GP73 antibody (I), or alanine tolerance test (ATT, J) in mice 24 h after rmGP73 injection (n=6 in each group). (K) Glycogen levels in mice 15 min after rmGP73 or PBS injection (n=3 in each group). (L) Schematic representation of the metabolic process responsible for GP73-induced hyperglycemia. The data are shown as the means ± SEM. **, P < 0.01; and ***, P < 0.001; one-way or two-way ANOVA followed by Bonferroni’s post hoc test.

Hyperglycemia is caused either by impaired glucose clearance and/or excess glucose production through glycogenolysis or gluconeogenesis. To determine which of these processes caused GP73-induced hyperglycemia, we performed an insulin tolerance test (ITT). There were no significant differences in mice challenged with rmGP73 (Figure 2F). However, when we assessed insulin resistance with the glucose tolerance test (GTT), we observed that a single dose of rmGP73 severely impaired glucose clearance (Figure 2G). The pyruvate tolerance test (PTT) indicated that the conversion of pyruvate to glucose was greatly enhanced after GP73 challenge in overnight-fasted, glycogen-depleted mice (Figure 2H) and that this effect was completely blocked by anti-GP73 treatment (Figure 2I). GP73 elevation also stimulated alanine-driven gluconeogenesis in a glycogen-depleted state (Figure 2J). Notably, no significant difference was found in hepatic glycogen content between rmGP73- and PBS-injected mice (Figure 2K), suggesting that glycogenolysis was not affected by GP73. Therefore, increase in circulating GP73 levels may elevate fasting blood glucose primarily by potentiating endogenous glucose production and reducing glucose clearance (Figure 2L).

### Circulating GP73 traffics to the liver and kidney and binds to the hepatocyte and renal cell surface

To examine the primary target of circulating GP73, rmGP73 labeled with Cy-7 (rmGP73-Cy7) was injected i.v. into mice, and an in vitro imaging system (IVIS) was used to identify sites of accumulation. An equivalent amount of free Cy7 was used as a control. In contrast to free Cy7, rmGP73-Cy7 trafficked primarily to the liver and kidney (Figure 3A) and exhibited higher accumulation in these two tissues 30 min after injection (Figures 3B-C). Ex vivo imaging 19 h post-administration revealed that the fluorescence signal in the liver was 20% of the signal at 1 h post-administration (Figures 3D and S2). No signal was detected in the other major organs 19 h post-rmGP73-Cy7 injection (Figure S2).

**Fig. 3.**
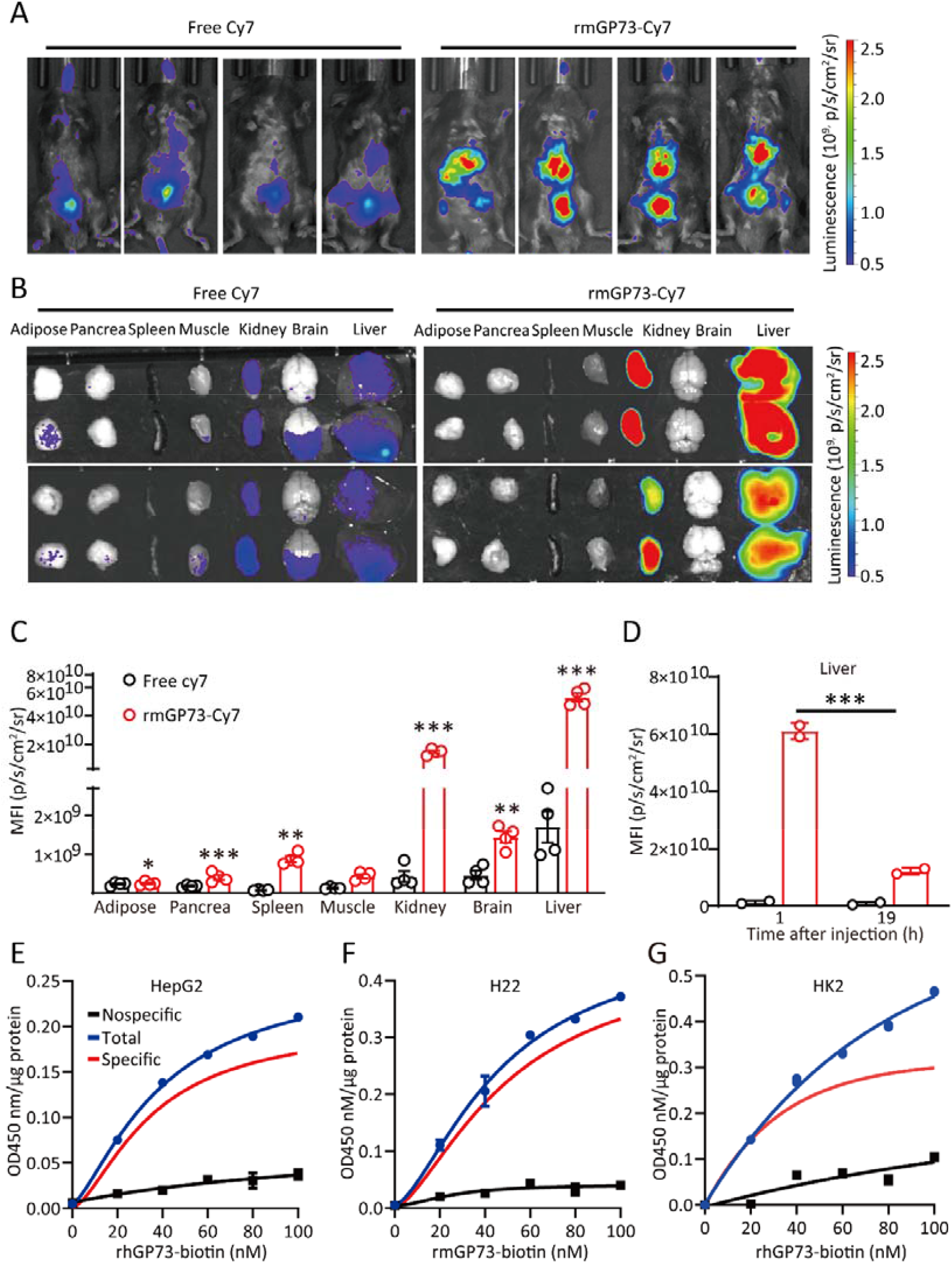
Circulating GP73 traffics to the liver and kidney and binds to the hepatocytes and renal surface. (A) In vivo imaging of live anesthetized mice was performed 30 min after i.v. injection of rmGP73-Cy7 or free Cy7 (n=4). (B) In vivo imaging of various organs from mice was performed 30 min after rmGP73-Cy7 or free Cy7 injection. Two representative images are shown. (C) Tissue GP73 accumulation in B was measured as the photon intensity. *, P < 0.05; **, P < 0.01; and ***, P < 0.001; two-tailed Student’s t-tests. (D) Liver GP73 accumulation in mice 1 h or 19 h after rmGP73-Cy7 or free Cy7 injection was measured as the photon intensity. ***, P < 0.001; one-way ANOVA followed by Bonferroni’s post hoc test. (E-G) The level of biotin on the hepatocyte surface upon incubation of HepG2 (E), H22 (F), and HK2(G) cells with increasing concentrations of rhGP73-biotin or rmGP73-biotin with (non-specific binding) or without (total binding) 100-fold excess recombinant GP73 in the medium was measured using a colorimetric assay. Specific binding (shown in red) was calculated as the difference between the two curves. Cell-based studies were performed independently at least three times with comparable results. The data are presented as the means ± SEMs.

To further examine the specific binding of GP73 to hepatocytes, we incubated mouse and human hepatocytes with increasing concentrations of rmGP73-biotin conjugate (rmGP73-biotin) or rhGP73-biotin conjugate (rhGP73-biotin). The cells were washed with PBS, and the relative level of biotin on the hepatocyte surface was measured. GP73 bound to the surface of human and mouse hepatocytes in a dose-dependent and saturable manner (Figure 3E-F). Similar binding results were observed for renal HK2 cells (Figure 3G). In particular, addition of 100-fold excess unconjugated GP73 abolished the binding (Figure 3E-G). These results suggest that circulating GP73 traffics and binds to the cell surface of the liver and kidney in a saturable and competitive manner.

### GP73 stimulates gluconeogenesis and induces insulin resistance

We then directly assessed the effect of GP73 on glucose production in isolated primary mouse hepatocytes (PMHs). Treatment of PMHs isolated from overnight-fasted mice with rmGP73 promoted HGP in a dose- and time-dependent manner (Figures 4A and S3A). A similar result was observed in rat primary hepatocytes treated with recombinant rat GP73 (rrGP73; Figure S3B). In particular, GP73-specific antibody blocked GP73-mediated hepatocyte glucose release, while a non-specific control antibody had no effect (Figure 4B). Additionally, a competitive antagonist of cyclic AMP (cAMP) binding to PKA, cAMPS-Rp, blocked GP73-mediated hepatocyte glucose release in a similar manner as glucagon (Figure S3C). The addition of rmGP73 also increased the expression of key gluconeogenic enzymes, including *Pcx, Pck1*, and *G6pc* (Figure 4C). Similar to what we observed for hepatocyte glucose release, treatment of PMHs with rmGP73 significantly increased intracellular cAMP levels and the kinase activity of PKA (Figure 4D-E). Phosphorylation of the PKA-C-α subunit and PKA substrate levels in hepatocytes and renal cells were also significantly increased after the addition of rhGP73 (Figure 4F-G). Notably, increased levels of cAMP response element-binding (CREB) phosphorylation and *G6pc* expression were observed in liver tissues from mice exposed to a single dose of rmGP73 (Figure 4H and S3D).

**Fig. 4.**
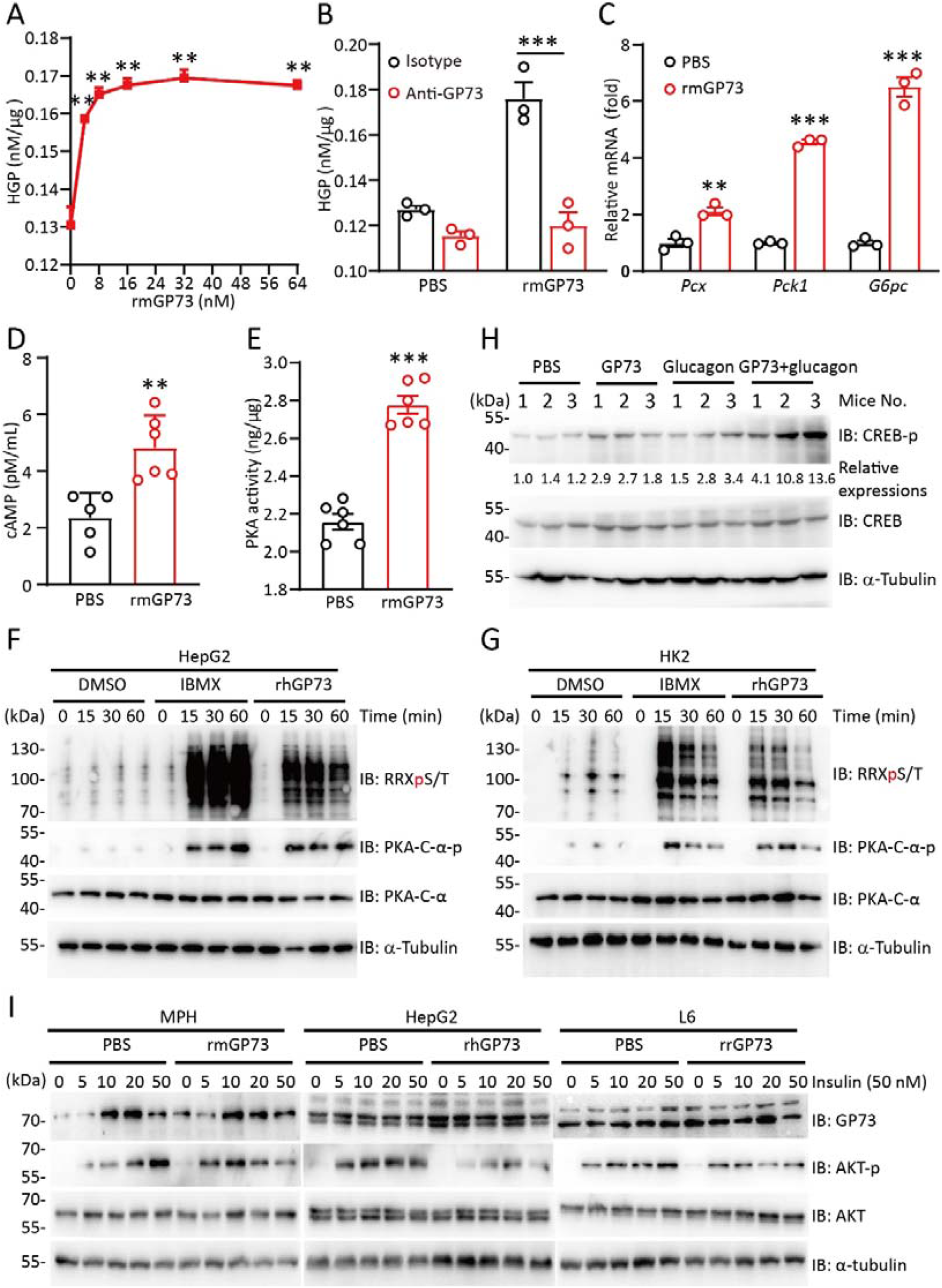
Circulating GP73 stimulates gluconeogenesis and induces insulin resistance. (A) Glucose production in PMHs treated with rmGP73 at the indicated concentrations for 2 h. (B) Glucose production in PMHs treated with 32 nM rmGP73 with or without anti-GP73 antibody for 2 h. (C-E) Gluconeogenesis gene expression (C), intracellular cAMP accumulation (D), and hepatocyte PKA activity (E) in PMHs treated with 32 nM rmGP73 for 2 h. (F-G) Immunoblotting analysis of phosphorylated PKA C subunit (PKA-C-α-p) levels and substrate (phosphoRRXT*T*-PKA substrate, RRXpS/T) levels in HepG2 (F) or HK2 (G) cells treated with 32 nM rmGP73 or IBMX for the indicated times. (H) Immunoblotting analysis of phosphorylated CREB (CREB-p) in the livers of mice treated with a single dose of i.v. rmGP73, glucagon, or both for 15 min (n = 3 in each group). (I) Immunoblotting analysis of phosphorylated AKT (AKT-p) in PMHs, HepG2, or L6 cells treated with the indicated concentrations of insulin in the presence or absence of GP73. Cell-based studies were performed independently at least three times with comparable results. The data are presented as the means ± SEMs. **, P < 0.01; and ***, P < 0.001; one-way or two-way ANOVA followed by Bonferroni’s post hoc test for A-C and two-tailed Student’s t-tests for D-E.

We next examined the effect of GP73 on insulin sensitivity. GP73 treatment tended to decrease insulin-stimulated phosphorylation of AKT at Ser473 in PMHs, HepG2 cells, and mouse myotube L6 cells (Figure 4I). Together, these results suggest that GP73 regulates hepatocyte glucose production and peripheral insulin sensitivity.

### GP73 induces a drastic remodeling of the PKA kinase hub

To determine the full extent of the hepatic signaling effect of circulating GP73 at the cellular level in the absence of systemic factors that may exert compensatory effects, we performed global phosphoproteomics analysis in PMHs treated with PBS, rmGP73, or glucagon in vitro. A total of 183 and 229 phosphosites were identified in the GP73-treated and glucagon-treated samples, respectively. The two samples shared approximately 27% of the upregulated phosphosites and 6% of the downregulated phosphosites (Figure 5A). Of the top 30 phosphosites that were highly upregulated by GP73, over 75% were also affected by glucagon (Figures 5B), indicating that these two proteins have overlapping functions at the signaling level. Among the potentially functional phosphosites regulated by GP73, ser1588 of inositol triphosphate receptor-1 (Itpr1, also known as InsP3R-I) attracted our attention (Figure 5C). This protein is a target of PKA and plays a crucial role in glucagon-stimulated hepatic gluconeogenesis. Other significantly regulated proteins by GP73 known to be involved in gluconeogenesis signaling included PHKA2^S729^, GNMT^Y221^, SLC16A1^S491^, MLX^S45^, BAD^S155^, CTNNB1^S552^, GPX1^S7^, CYP2E1^S129^, and HCFC1^S1516^ (Figure 5C).

**Fig. 5.**
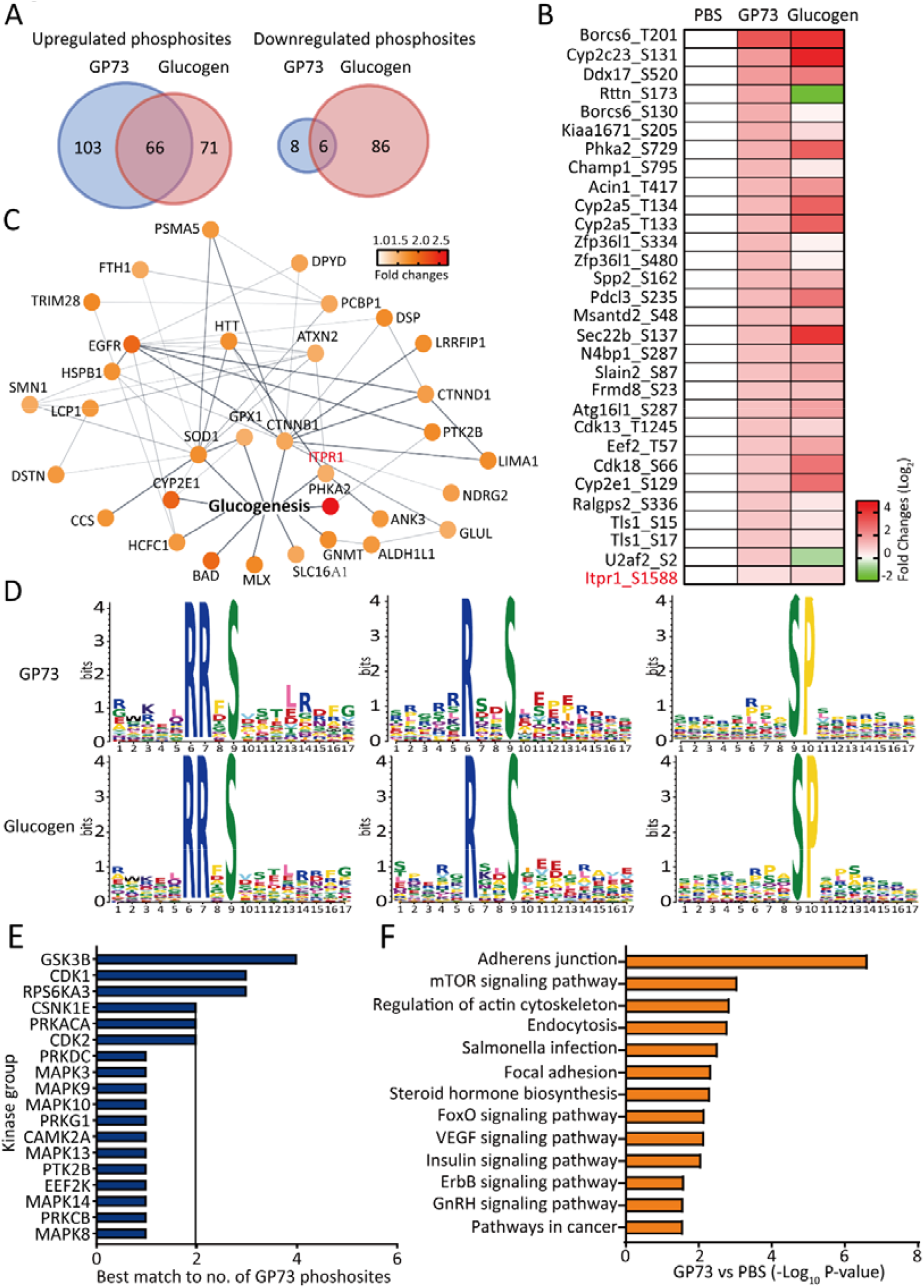
GP73 induces drastic remodeling of the PKA kinase hub. (A) Overlapping numbers of significantly upregulated phosphopeptides and downregulated phosphopeptides in PMHs treated with rmGP73 (64 nM) or glucagon (3 μM) for 1 h. Localization probability ≥0.75 and fold-change ≥1.5 or fold-change ≤ 0.67 were declared significant. Significantly regulated phosphopeptides by GP73 are shown in blue, and significantly regulated phosphopeptides by glucagon are shown in red. (B) Heatmap of the top 30 highly upregulated phosphosites in PMHs treated with rmGP73. (C) Network analysis of proteins involved in the glucogenesis signaling pathway in PMHs treated with rmGP73. Each of the phosphosites are color-coded based on the fold changes. Circular shapes show each protein that is upregulated in GP73-treated cells compared to PBS-treated cells. Lines indicate protein-protein interactions curated from databases of experimentally defined kinase-substrate relationships (STRING, confidence >0.7). (D-E) Specific kinase substrate motifs (D) or distribution of matching kinases according to the phosphoproteomics data from the rmGP73-treated sample (P <0.05) using MoMo (http://meme-suite.org/tools/momo) and Kinase Enrichment Analysis 2 (KEA2). (F) KEGG enriched pathway analysis of significantly regulated phosphopeptides in PMHs treated with rmGP73 (P <0.05) using DAVID Bioinformatics Resources 6.8. The bar plot shows significantly dysregulated pathways, and Fisher’s exact test P values are shown on the x-axis.

To investigate the mechanisms responsible for the phosphoproteome changes, kinase substrate motifs were extracted, and a clear shift towards PKA kinase activation following GP73 or glucagon stimulation was identified (Figure 5D). Other substrate motifs of key kinases previously implicated in the regulation of gluconeogenesis and insulin resistance, including GSK3B, PRKG1, PTK2B, and CaMK2A, were also dysregulated, which further explains the ability of GP73 to affect glucose production and insulin sensitivity (Figure 5E and S4A). Additional enriched kinase motifs following GP73 stimulation included mitogen-activated kinase (MAPK) 3/8/9/10/13/14 and cyclin-dependent kinase (CDK)1/2 (Figure 5E). Global kinase enrichment analysis following GP73 stimulation revealed that GP73 induced a drastic remodeling of PKA hubs in a similar manner as glucagon (Figure S5A-B). Pathway analysis of the phosphosites regulated by GP73 revealed pathways related to cell-cell adhesion, cell endocytosis, mRNA processing, gene transcription, and cell cycle regulation (Figure S4B). At the signaling level, the mTOR pathway, FoxO pathway, and insulin pathway were positively enriched in GP73-treated hepatocytes (Figure 5F). Therefore, GP73 induces a drastic remodeling of the PKA kinase hub and multiple signaling pathways.

### GP73 secretion is induced from multiple tissues upon fasting

Given the glucogenic roles of GP73 in glucose homeostasis, we hypothesized that GP73 levels were regulated in response to fasting signals. Indeed, fasting in humans and mice increased plasma GP73 levels (Figure 6A-B). Since intracellular GP73 overexpression led to the release of GP73 into the extracellular space (Figure S6A-B), we examined and compared GP73 expression in metabolically important organs isolated from ad libitum-fed and fasted mice to delineate the source of GP73 under fasting conditions. After 24 h of fasting, we found a strong upregulation of GP73 mRNA expression in the heart, liver, kidney, white adipose tissue (WAT), and pancreas (Figure 6C). The upregulation of GP73 in the liver persisted for 48 h of fasting then dropped to levels lower than the fed and refed states (Figure 6D). Notably, the upregulated GP73 in the kidney was progressively elevated with prolonged fasting times compared to its levels in the fed and refed states (Figure 6D). Consistent with this finding, the intensity of GP73 immunofluorescence staining in liver, WAT, and pancreatic tissues was significantly increased upon 24 h of fasting (Figure 6E). GP73 secretion from cultured HepG2 cells was also promoted in response to the fasting-related signal forskolin (FSK) (Figure 6F).

**Fig. 6.**
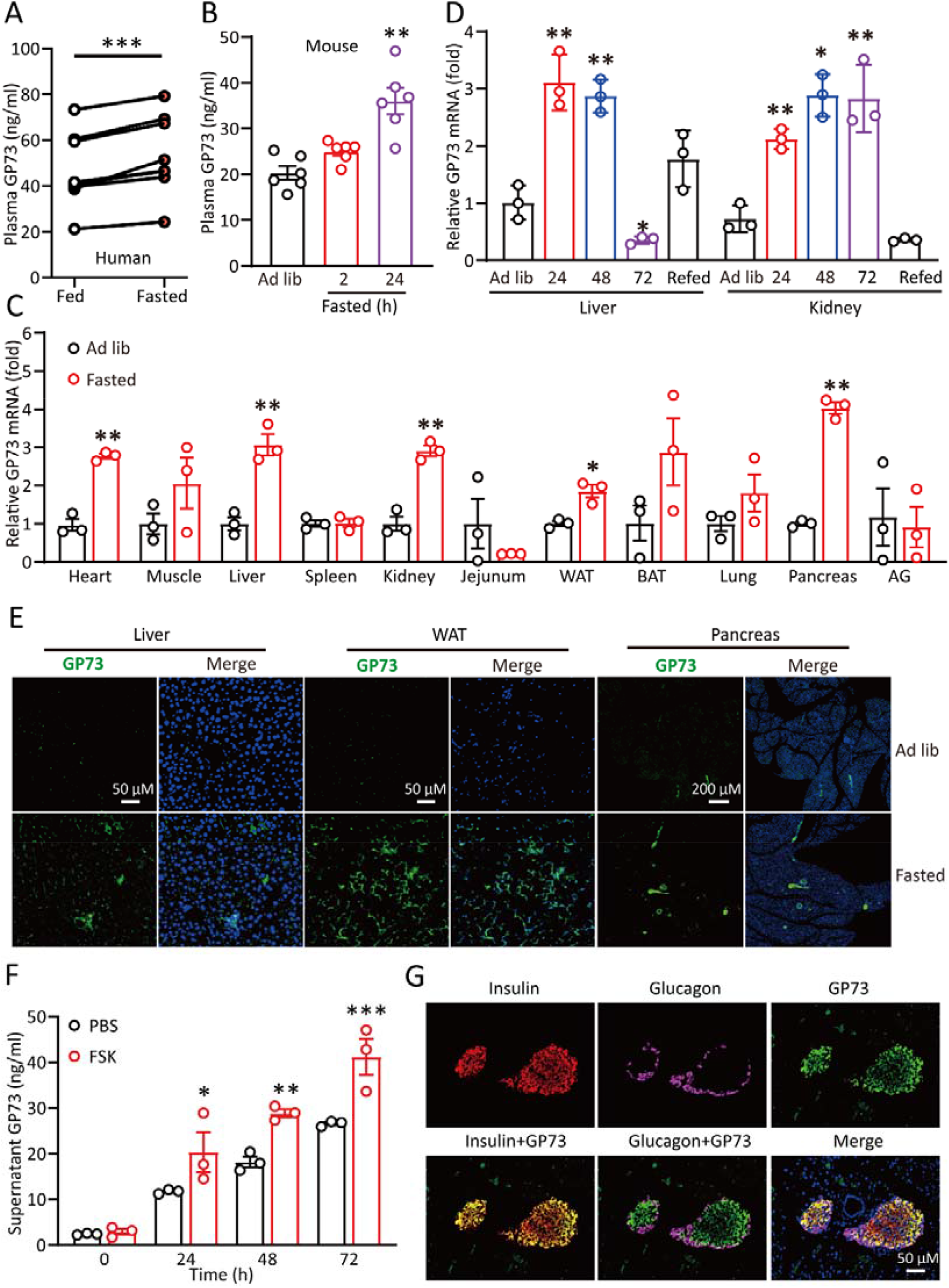
GP73 secretion is induced from multiple tissues upon fasting. (A) GP73 levels in blood samples obtained from humans following breakfast or following an overnight fast (n=7). **, P < 0.01, and ***, P < 0.01; two-tailed Student’s t-tests. (B) Plasma GP73 levels in mice fed ad libitum or fasted for 24 h (n=6). (C) QRT-PCR analysis of GP73 mRNA expression in various organs from mice fed ad libitum or fasted for 24 h (white adipose tissue, WAT; brown adipose tissue, BAT; adrenal gland, AG; n=3). *, P < 0.05, and **, P < 0.01; two-tailed Student’s t-tests. (D) QRT-PCR analysis of GP73 mRNA expression in the livers or kidneys of mice fasted for the indicated times (n=3). (E) Representative confocal immunofluorescence images of GP73 staining in liver, WAT, and pancreatic sections from mice fed ad libitum or fasted for 24 h. Green represents GP73, blue represents nucleus. (F) Supernatant GP73 levels in HepG2 cells treated with FSK (20 µM) for the indicated times. (G) Representative immunofluorescence staining of GP73 (green), glucagon (purple), and insulin (red) in pancreatic sections from mice fed ad libitum or fasted for 24 h. Blue represents nucleus. Cell-based studies were performed independently at least three times with comparable results. The data are presented as the means ± SEMs. *, P < 0.05; **, P < 0.01; and ***, P < 0.001; one-way ANOVA followed by Bonferroni’s post hoc test for D, F.

To further identify the pancreatic cell type that produced GP73 in response to fasting, we examined GP73 staining in islets isolated from fasted mice using triple immunofluorescence staining for glucagon (a marker for α cells), insulin (a marker for β cells) and GP73. Specifically, GP73 primarily colocalized with β cells (Figures 6G and S6C). Taken together, GP73 secretion is induced from multiple cell types upon fasting.

### GP73 secretion is promoted under glucotoxicity conditions associated with SARS-CoV-2 infection and diabetes

Since excessive gluconeogenesis directly predisposes the host to abnormal glucose metabolism, we then want to examine the clinical association between GP73 expression and blood glucose levels in patients with COVID-19. Blood glucose levels in COVID-19 patients were higher than the reference population and were particularly higher in severe patients (Figure 7A and S7). The plasma concentrations of GP73 in COVID-19 patients were strikingly correlated with blood glucose levels (Figure 7B). We next measured the plasma concentrations of GP73 in 100 healthy and 190 diabetic individuals (Table 3). Diabetic patients had significantly higher plasma GP73 levels than age- and sex-matched healthy subjects (Figure 7C). The plasma concentrations of GP73 correlated with average blood glucose levels (glycated A1c, HbA1c) (Figure 7D).

**Table 3.** Clinical characteristics of diabetics and reference controls.

| Parameter | Healthy<br>(n=100) | Diabetics<br>(n=190) | P value |
| --- | --- | --- | --- |
| **Clinical characteristics** |  |  |  |
| Age (y) | 58 ± 15 | 62 ± 11 | 0.0102 |
| Male (n) | 59 | 102 |  |
| Female (n) | 41 | 88 |  |
| **Laboratory examination** |  |  |  |
| Leukocytes (10<sup>9</sup>/L) | 5.13 ± 2.29 | 7.82 ± 3.39 | < 0.0001 |
| Lymphocytes (10<sup>9</sup>/L) | 1.31 ± 0.45 | 1.74 ± 0.63 | < 0.0001 |
| Neutrophils (10<sup>9</sup>/L) | 3.05 ± 1.56 | 5.43 ± 3.40 | < 0.0001 |
| Monocytes (10<sup>9</sup>/L) | 0.41 ± 0.21 | 0.49 ± 0.24 | 0.0052 |
| Platelets (10<sup>9</sup>/L) | 215.56 ± 76.67 | 230.88 ± 54.29 | 0.04959 |
| CRP (mg/L) | 5.41 ± 2.65 | 10.71 ± 8.72 | < 0.0001 |
| ALT (40 U/L) | 18.61 ± 9.58 | 32.03 ± 13.39 | < 0.0001 |
| AST (40 U/L) | 16.16 ± 4.80 | 26.41 ± 14.26 | < 0.0001 |
| BG (mmol/L) | 88.21 ± 7.73 | 177.66 ± 63.00 | < 0.0001 |
| Abbreviations: CRP, C-reactive protein; ALT, alanine transaminase; AST, Aspartate transaminase; BG, blood glucose.<br>P values of differences between the two groups were calculated using the Mann-Whitney U test for continuous variables. |  |  |  |

**Fig. 7.**
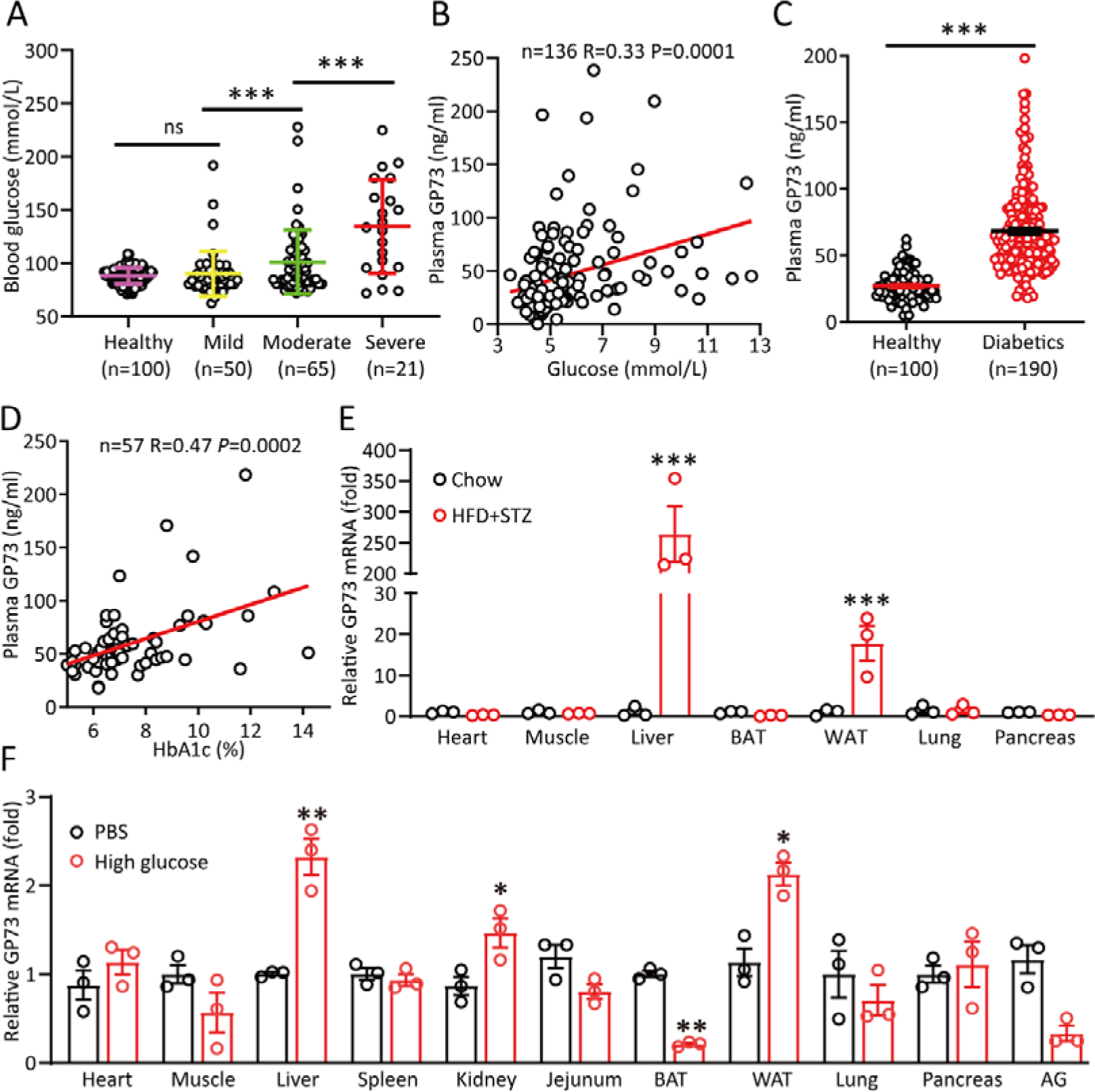
GP73 secretion is promoted under glucotoxicity conditions associated with SARS-CoV-2 infection and diabetes. (A) Blood glucose levels in healthy controls (n=100) and COVID-19 patients with different disease severities. ns, no significant; and ***, P < 0.001; Mann-Whitney U test. (B) Correlation analysis between plasma GP73 levels and glucose levels in COVID-19 patients using Spearman’s non-parametric test (n = 136; R =0.33, p=0.0001). (C) Plasma GP73 levels in healthy (n=100) and diabetic patients (n=190). ***P < 0.001; Mann-Whitney U test. (D) Correlation analysis between plasma GP73 levels and HbA1c levels in diabetic patients using Spearman’s non-parametric test. (E-F) QRT-PCR analysis of GP73 mRNA expression in various mouse organs from mice fed an HFD and injected with STZ at week 8 or a regular diet (chow; E) or mice at 30 min after i.v. injection of PBS or high glucose (0.5 g/mg; F) (n=3 in each group). The data are presented as the means ± SEMs. *, P < 0.05; **, P < 0.01; and ***, P < 0.001; two-tailed Student’s t-tests.

To assess whether hyperglycemia is a potential driving force in the upregulation of GP73 expression, we assessed GP73 mRNA expression profiles in various tissues from mice fed a regular diet (chow) or HFD plus streptozotocin (STZ)-induced T2D mice. We found a strong upregulation of GP73 mRNA expression in WAT and liver (Figure 7E). The upregulation in liver tissue was notably striking, indicating that the liver is a major source of plasma GP73 under glucotoxicity conditions. To continue our investigation, we measured GP73 mRNA expression in mice injected with a high dose of glucose (0.5 g/kg) in the fasted state. We found a strong upregulation of GP73 expression in WAT, kidney, and liver (Figure 7F). Therefore, the pathophysiological relevance of GP73 to hyperglycemia associated with SARS-CoV-2 infection and diabetes was established.

### GP73 is essential for SARS-CoV-2-induced gluconeogenesis exaggeration

After confirming the roles of GP73 as a glucogenic hormone, we hypothesized that SARS-CoV-2-induced hyperglycemia might result from excessive gluconeogenesis triggered by the upregulation of GP73 expression. To test this hypothesis, we assessed the impact of SARS-CoV-2 infection on gluconeogenesis metabolism in Huh-7 cells. Notably, SARS-CoV-2 infection led to significantly increased expression of key glucogenic genes and glucose production (Figure 8A-B). PKA activity was also significantly increased in infected hepatocytes (Figure 8C). Therefore, SARS-CoV-2 infection induced enhanced liver gluconeogenesis by activation of PKA signaling. We then proceeded to investigate whether GP73 is involved in this effect. Indeed, PKA activation by SARS-CoV-2 infection was significantly blocked by GP73-specific antibody (Figure 8C). Meanwhile, the stimulatory effects of SARS-CoV-2 on the induction of glucogenic gene expression and hepatocyte glucose production were also markedly reduced by anti-GP73 antibody treatment (Figure 8A-B), similar to the effect of GP73 depletion (Figure 8D-E).

**Fig. 8.**
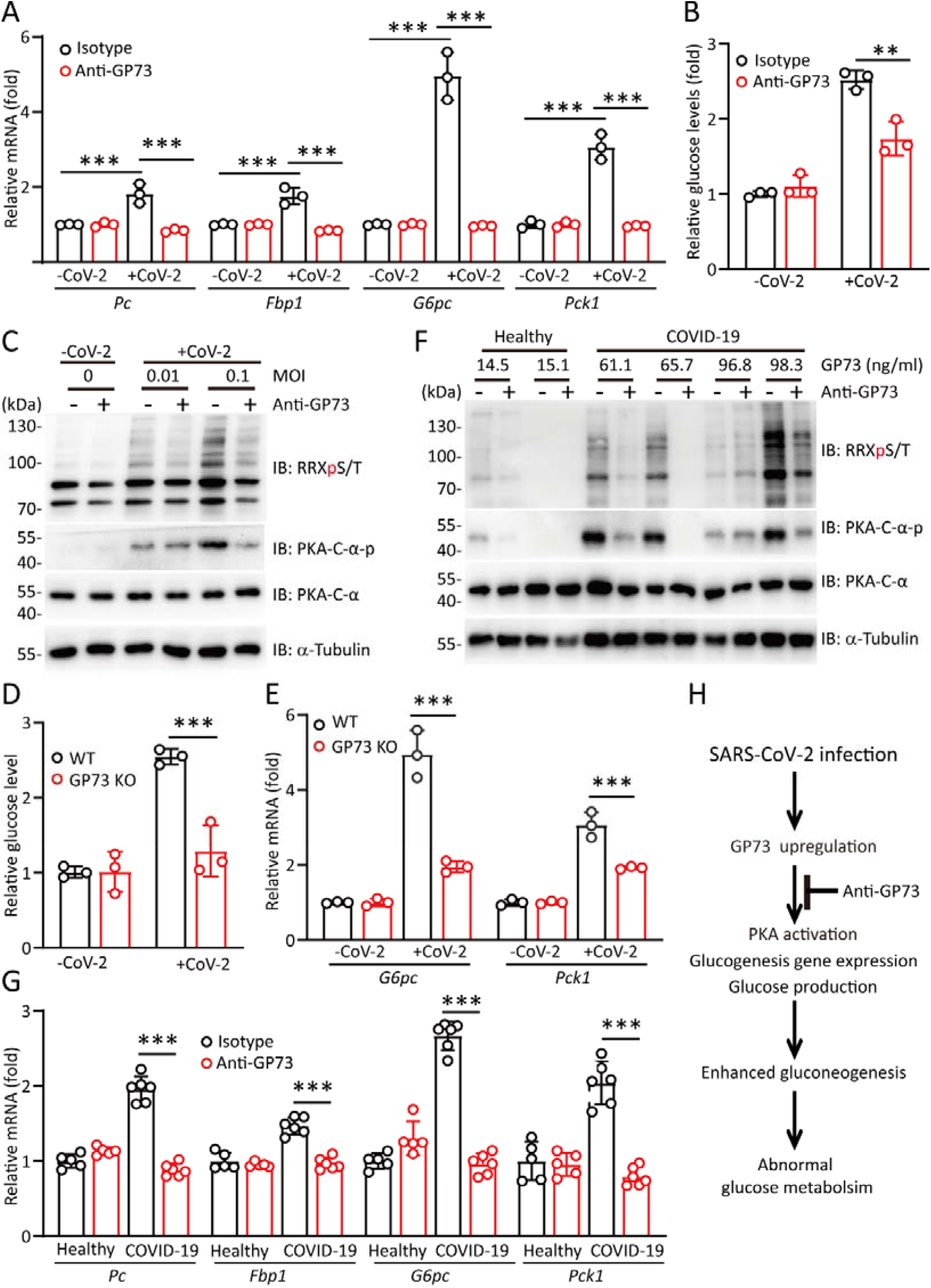
GP73 is essential for SARS-CoV-2-induced exaggeration of gluconeogenesis metabolism. (A) QRT-PCR analysis of glucogenic gene expression in Huh-7 cells infected with SARS-CoV-2 (MOI 0.1) for 24 h with or without GP73 antibody (1 µg/mL). (B) Glucose levels in Huh-7 cells infected with SARS-CoV-2 (MOI 0.1) for 24 h in glucose-free medium with or without GP73 antibody (1 µg/mL). (C) Immunoblotting analysis of PKA-C-α phosphorylation and substrate levels in Huh-7 cells infected with SARS-CoV-2 (MOI 0.1) for 24 h with or without anti-GP73 antibody. (D) Glucose levels in WT and GP73 KO-1 Huh-7 cells infected with SARS-CoV-2 (MOI 0.1) for 24 h in glucose-free medium. (E) QRT-PCR analysis of glucogenic gene expression in WT and GP73 KO-1 cells infected with SARS-CoV-2 (MOI 0.1) for 24 h. (F-G) PKA enzymatic activity (F) or glucogenic gene expression (G) in Huh-7 cells cultured with serum from healthy controls or COVID-19 patients in the presence or absence of anti-GP73 antibody. (H) Schematic representation of the involvement of upregulated GP73 in SARS-CoV-2-induced gluconeogenesis exaggeration. Cell-based studies were performed independently at least three times with comparable results. The data are presented as the means ± SEMs. *, P < 0.05; **, P < 0.01; and ***, P < 0.001; one-way ANOVA followed by Bonferroni’s post hoc test.

To further investigate whether the changes in serum GP73 were closely associated with gluconeogenesis levels, we obtained serum from COVID-19 patients with GP73 levels higher than 60 ng/ml and age-matched healthy individuals with GP73 levels lower than 20 ng/ml and added the samples to culture medium. Glucogenic gene expression and PKA enzymatic activity were more strongly induced in cells exposed to serum from COVID-19 patients than cells exposed to serum from healthy controls (Figure 8F-G). Treatment with an anti-GP73 antibody significantly blocked the PKA activation and upregulated glucogenic gene expression induced by serum from COVID-19 patients (Figure 8F-G). Therefore, we concluded that circulating GP73 was necessary for SARS-CoV-2-induced gluconeogenesis enhancement.

### GP73 blockade reduces gluconeogenesis and lowers hyperglycemia associated with diabetes

Since targeting GP73 is a therapeutic strategy for treating patients with this altered hormone during insulin resistance, the effect of GP73 antibody on excessive gluconeogenesis associated with diabetes was then explored. As expected, PKA enzymatic activity was more strongly induced in hepatocytes exposed to serum from diabetic patients than serum from healthy controls (Figure 9A). Specifically, treatment with an anti-GP73 antibody blocked PKA activation induced by serum from diabetic patients (Figure 9B).

**Fig. 9.**
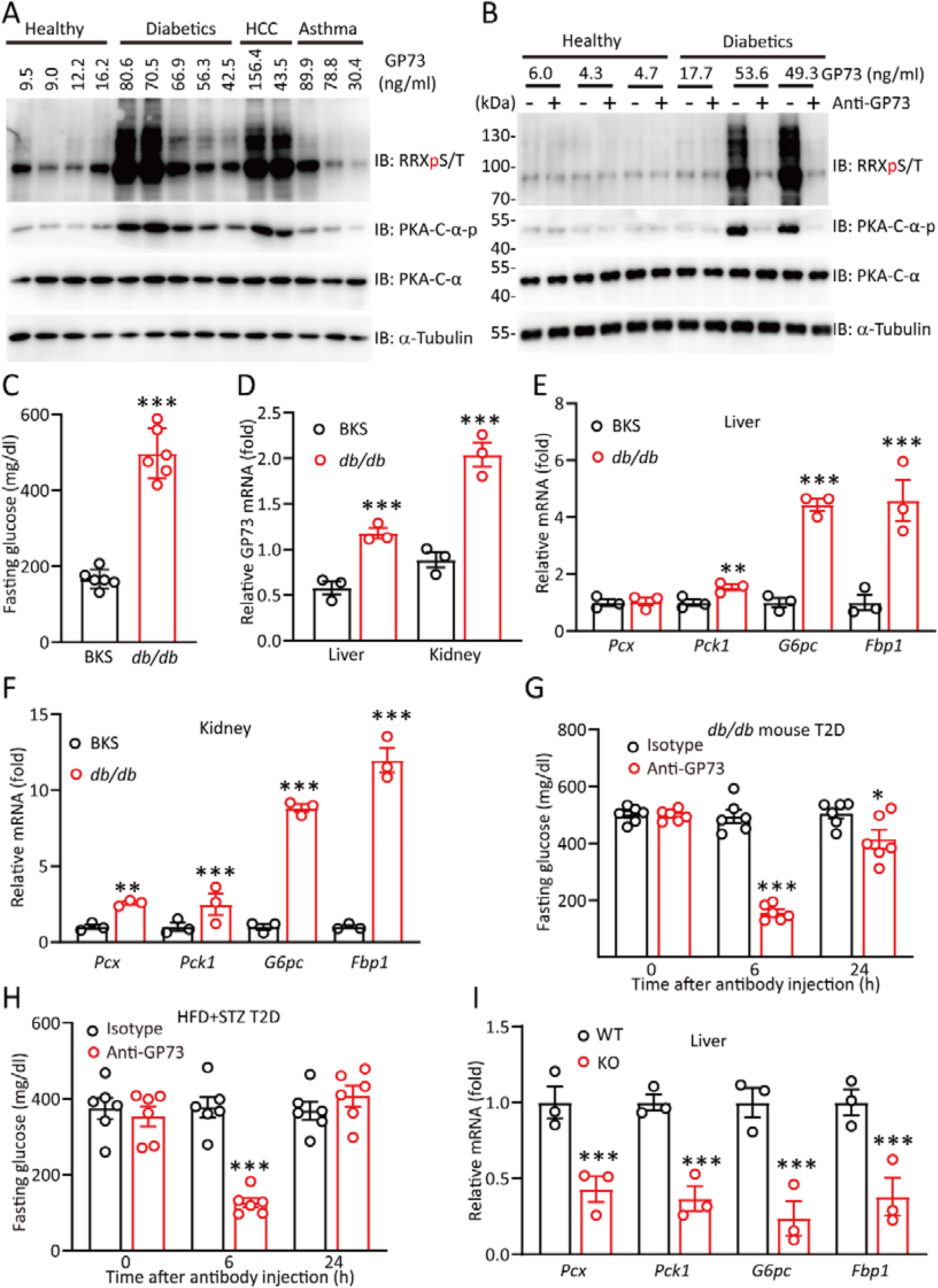
GP73 blockade reduces gluconeogenesis and lowers hyperglycemia associated with diabetes. (A-B) Immunoblotting analysis of PKA-C-α phosphorylation and substrate levels in HepG2 cells cultured with serum from healthy subjects, diabetic subjects, hepatocellular carcinoma (HCC) patients, asthmatic subjects (A) or serum from healthy or diabetic subjects with or without anti-GP73 antibody (B). (C-F) Fasting glucose level (C; n=6 in each group), hepatic and renal GP73 mRNA expression (D; n = 3 in each group), hepatic glucogenic gene expression (E; n = 3 in each group), and renal glucogenic gene expression (F; n = 3 in each group) in BKS and db/db mice. (G-H) Fasting blood glucose levels in db/db mice (G) or HFD+STZ-induced T2D mice (H) at the indicated times after anti-GP73 treatment (n=6 in each group). (I) QRT-PCR analysis of glucogenic gene expression in the livers of WT and GP73 KO adult mice fasted for 6 h (n=3 in each group). The data are the means ± SEM. **, P < 0.01, and ***, P < 0.001; two-tailed Student’s t-tests for C and one-way ANOVA followed by Bonferroni’s post hoc test for D-I.

To further confirm this therapeutic effect, we used *db/db* T2D mice, which exhibit severe fasting hyperglycemia (Figure 9C), increased hepatic and renal GP73 expression (Figure 9D), and upregulated glucogenic gene expression (Figure 9E-F). Fasting blood glucose levels were significantly decreased in GP73 antibody-treated mice compared to IgG-treated mice 6 h after administration (Figure 9G). A similar result was obtained in the HFD+STZ-induced T2D mice model (Figure 9H). Accordingly, hepatic and renal gluconeogenic gene expression was significantly lower in GP73 KO mice than WT mice (Figures 9I and S8). All these results indicate that GP73 blockade has a profound glucose-lowering effect that is primarily due to reduced gluconeogenesis.

## Discussion

Precise regulation of gluconeogenesis required for physiological adaptation to fasting and starvation occurs at multiple levels, including hormone secretion, gene transcription, posttranslational modification, and substrates flux(Romere et al., 2016; Zhang et al., 2018). In response to stimulation by external factors, the circulating blood levels of insulin, glucagon and glucocorticoids change, leading to subsequent changes in glucogenic pathways, regulation of gene expression and glucose production(Yabaluri and Bashyam, 2010). Here, we provide evidence linking circulating GP73 to gluconeogenesis. Consistent with the fact that hepatic glucose release is necessary during fasting, the circulating concentration of GP73 rises during fasting. Experimentally changing the circulating GP73 concentration via direct recombinant protein injection induced corresponding changes in fasting blood glucose levels. Mechanistically, secreted GP73 primarily targeted the liver and kidney to promote gluconeogenesis via activation of PKA signaling in endocrine and autocrine manners. By establishing a pathological role for elevated serum GP73 in excessive gluconeogenesis associated with SARS-COV-2 infection and diabetes, we hypothesize that the hyperglycemia during SARS-COV-2 infection or lifestyle-related diabetes might partly converge on the glucogenic property of GP73.

We assessed the direct actions of GP73 in the regulation of glucose production in mouse and rat primary hepatocytes in the absence of other counterregulatory hormones. GP73 was sufficient to cause cAMP accumulation, stimulate the enzymatic activity of PKA, increase the phosphorylation of CREB, and enhance the transcriptional expression of *Pcx, Pck1*, and *G6pc*. These results suggest that GP73 stimulates gluconeogenesis via the activation of PKA signaling through transcriptional regulation. In support of this, we observed a drastic remodeling of the PKA kinase hub following GP73 challenge using phosphoproteomics. Likewise, phosphosites that were upregulated by glucagon were also significantly enriched with substrate motifs for PKA kinase, indicating that glucagon and GP73 have overlapping functions at the signaling level. Of the unique kinase affected by GP73, protein kinase Cβ and p38 MAPK were identified, both of them contributes to hepatic steatosis and insulin resistance. Identification of the cell-surface receptor through which GP73 exerts its effects may provide more mechanistic information.

Epidemiological and experimental data indicate that SARS-CoV-2 infection is involved in the development of diabetes. The mechanisms underlying this phenomenon are complex and may include the promotion of inflammation, structural lung damage, and systemic effects. The present study reported that SARS-CoV-2 infection was associated with insulin resistance via pathways involved in endogenous glucose production. We demonstrated that enhanced gluconeogenic metabolism following SARS-CoV-2 infection was primarily dependent on GP73. Targeting GP73 may be a therapeutic strategy for the treatment of patients with altered levels of this hormone during SARS-CoV-2 infection.

We established a positive correlation between serum GP73 and blood glucose levels in patients with SARS-CoV-2 infection and diabetics, and observed increased GP73 expression in multiple tissues from mice subjected to high glucose treatment or under glucotoxic conditions. These results suggest that the persistent elevation of GP73 levels in patients with late-stage COVID-19 may be driven primarily by suboptimal blood glucose control rather than the infection per se. The exact drivers for the stimulation of GP73 expression in physiological and pathological states require further investigation. However, we hypothesized that the alterations in glucose metabolism following sudden onset of COVID-19 will persist even after the infection resolves. In fact, it has been observed that hyperglycemia persists for 3 years after recovery from SARS(Yang et al., 2010). Apart from the role of GP73 in promoting cell proliferation, tumor development, and metastasis(Wang and Wan, 2020), GP73 also represses the host innate immune response to promote RNA virus replication(Zhang et al., 2017). Therefore, long-term follow-up of infected patients is a priority.

In summary, our results provide convincing evidence that SARS-CoV-2 infection induces GP73 expression, which results in an exaggerated gluconeogenic response and may directly predispose the host to abnormal glucose metabolism. Our findings support the exciting possibility that neutralizing plasma GP73 may be an efficient approach for the treatment of infection- and lifestyle-related diabetes.

## Data Availability

Further information and requests for resources and reagents should be directed to and will be fulfilled by the Lead Contact Hui Zhong.

## Acknowledgements

We thank the National Key Research and Development Programme of China (grant no. 2018YFA0900800) and National Natural Science Foundation of China (grant nos. 31872715 32070755 81773205 and 82070595) for their support. Graphical abstract was created and exported with BioRender.com under a paid subscription.

## Author Contributions

X.L.Y., Q.C.F., Z.W.S., F.X.W., C.W.W and H.Z. designed the experiments. L.M.W., J.G., L.F.H, X.L.W, Q.G and Y.H.K. collected and analyzed the data. H.Y., J.G., X.P.Y., H.T.L., C.Q.L, F.Z. and Y.H.Z carried out mice assays. Y.Q.D. carried out authentic virus assys. L.M.W., H.L.L., C.J.W, D.Y.L, H.P.W, Y.M.P, L.X, J.L.L, X.M.Z and Q.L.Y. carried out cell lines experiments. C.W.W. and H.Z. analyzed the data and prepared the manuscript.

## Figure Legends

**Fig. S1.**
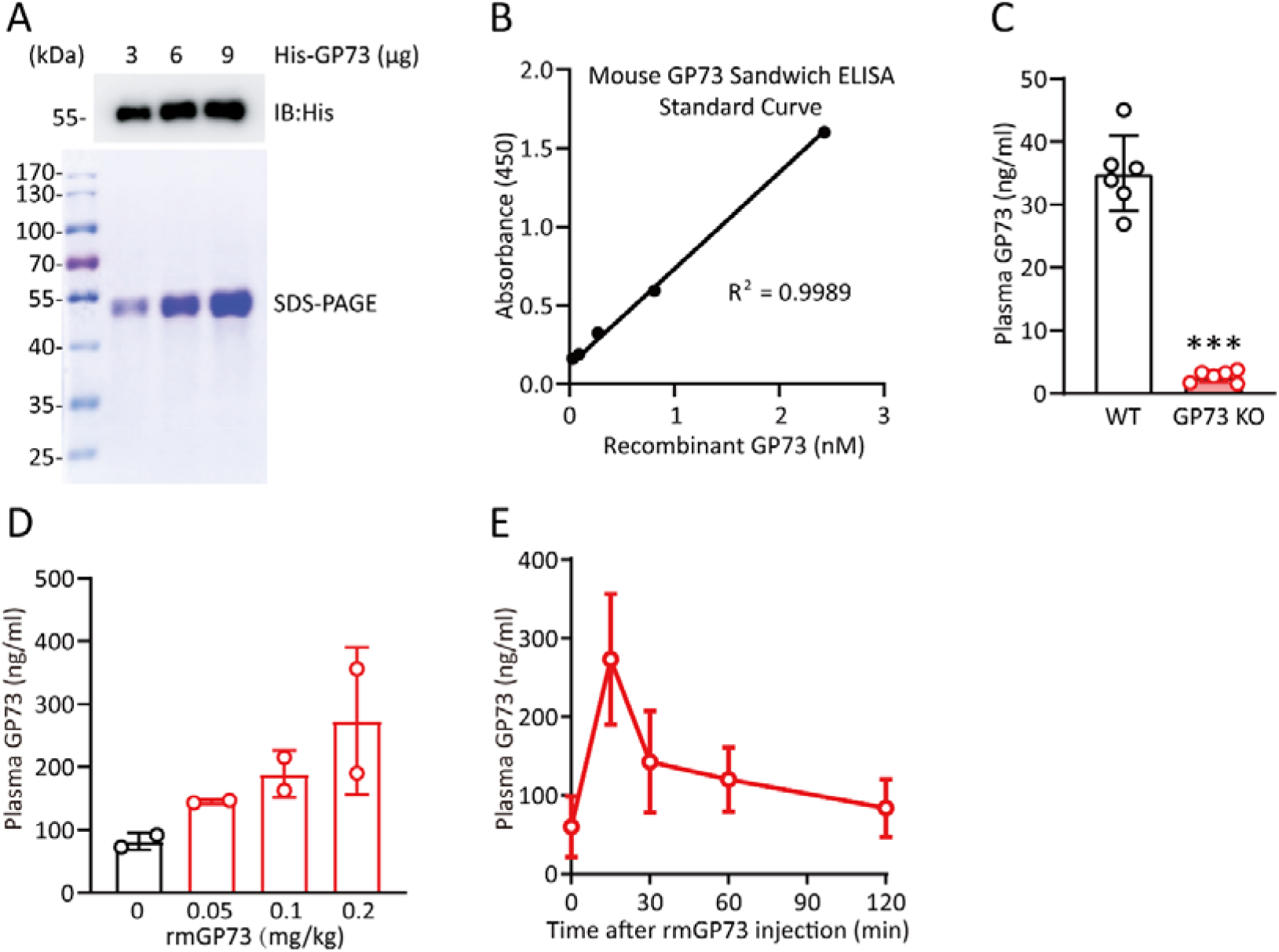
Increase in circulating GP73 elevates fasting blood glucose (related to Figure 2) (A) Immunoblotting analysis (upper lane) and SDS-PAGE (lower lane) of purified His-tagged rmGP73. (B) Mouse sandwich ELISA standard curve. (C) Plasma GP73 levels in WT and GP73 KO mice (n=6 in each group). ***P < 0.001; two-tailed Student’s t-tests. (D-E) Plasma GP73 levels in mice after rmGP73 injection at the indicated doses (D) for the indicated times (E) (n=2 in each group).

**Fig. S2.**
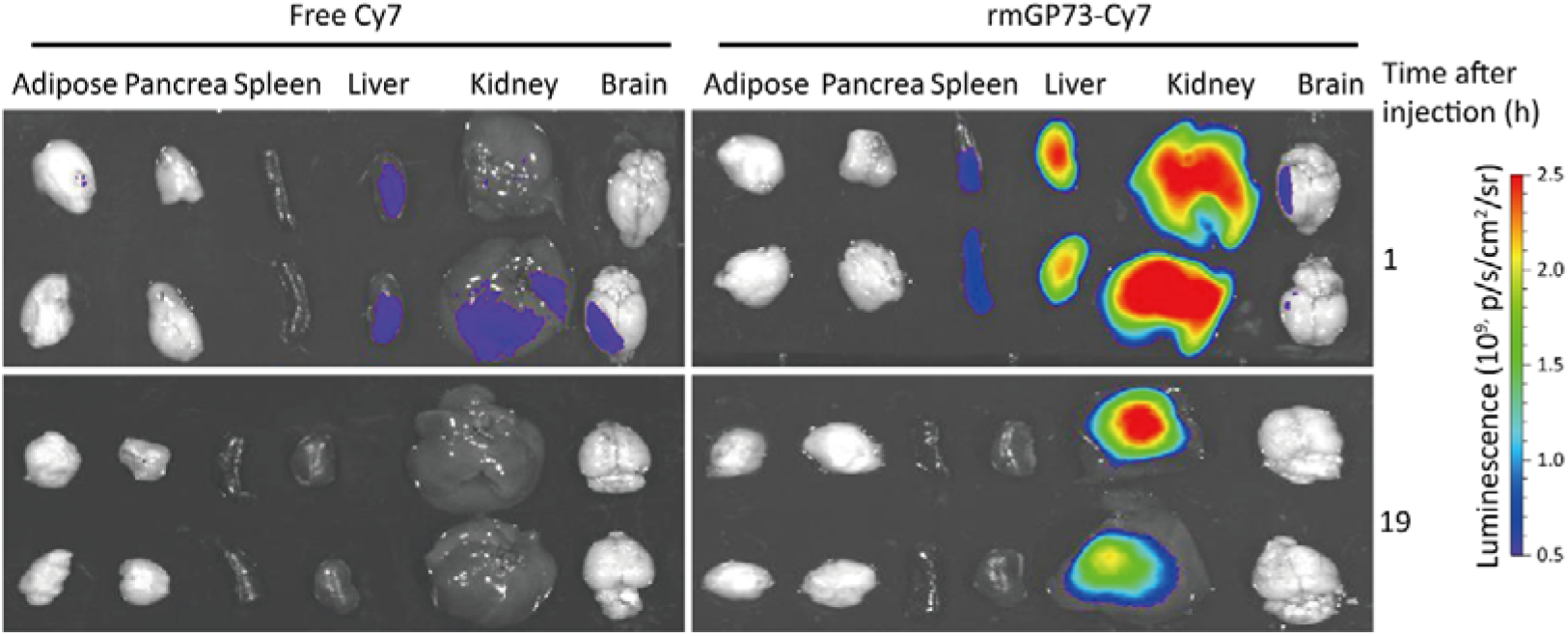
Circulating GP73 traffics to the liver and kidney and binds to the hepatocytes and renal surface (related to Figure 3) In vivo imaging of various organs from mice was performed 1 h or 19 h after rmGP73-Cy7 or free Cy7 injection. Two representative images are shown.

**Fig. S3.**
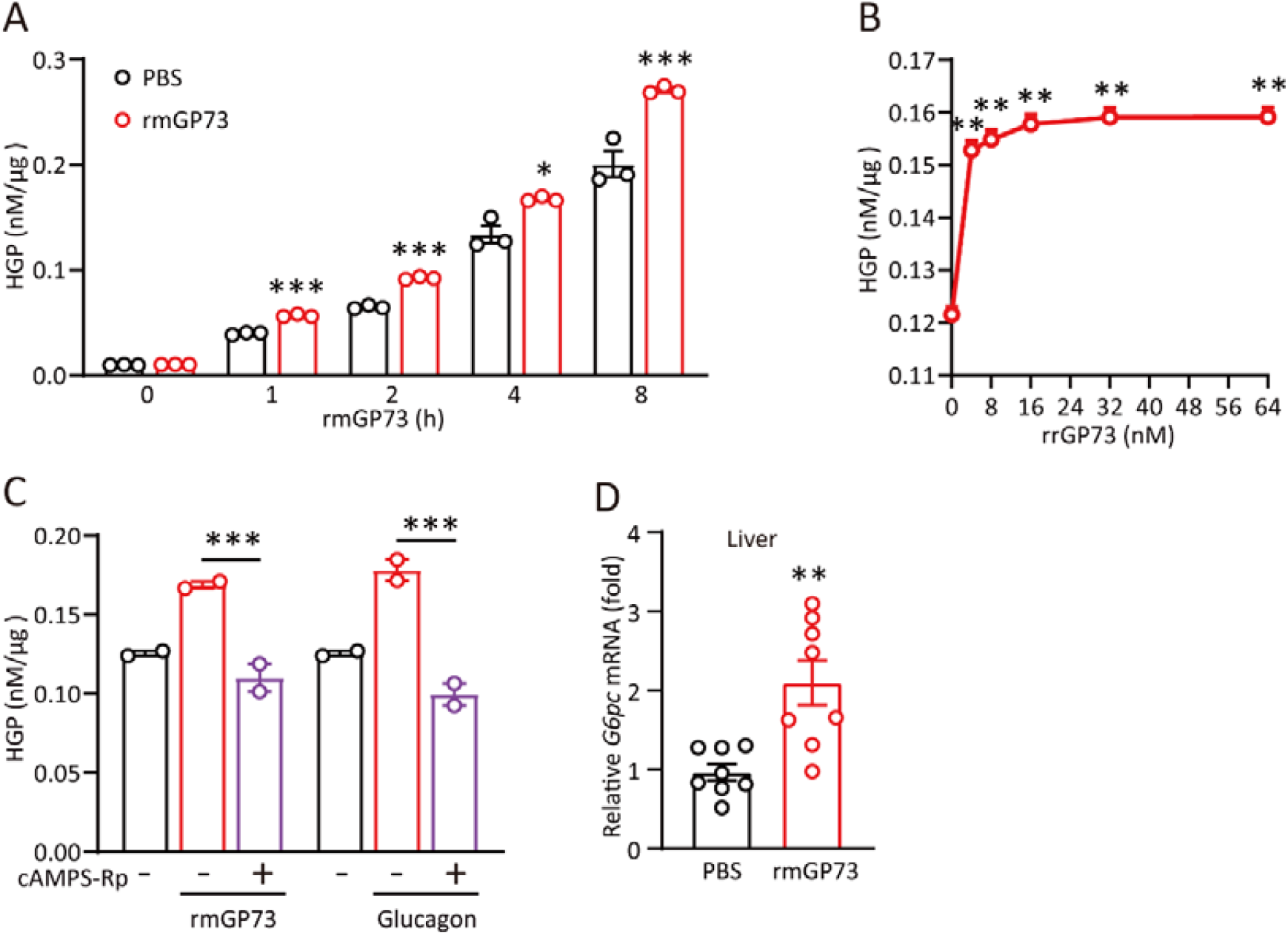
Circulating GP73 stimulates gluconeogenesis and induces insulin resistance (related to Figure 4) (A) Glucose production in PMHs treated with 32 nM rmGP73 for the indicated times. (B) Glucose production in primary rat hepatocytes treated with the indicated concentrations of rrGP73 for 2 h. (C) Glucose production in PMHs treated with 32 nM rmGP73 or glucagon in the presence or absence of cAMPS-Rp (200 μM). (D) QRT-PCR analysis of *G6pc* mRNA expression in the livers of mice treated with a single dose of i.v. rmGP73, glucagon, or both for 15 min (n = 7 or 8 in each group). Cell-based studies were performed independently at least three times with comparable results. The data are presented as the means ± SEMs. *, P < 0.05; **, P < 0.01; and ***, P < 0.001; one-way or two-way ANOVA followed by Bonferroni’s post hoc test for A-C and two-tailed Student’s t-tests for D.

**Fig. S4.**
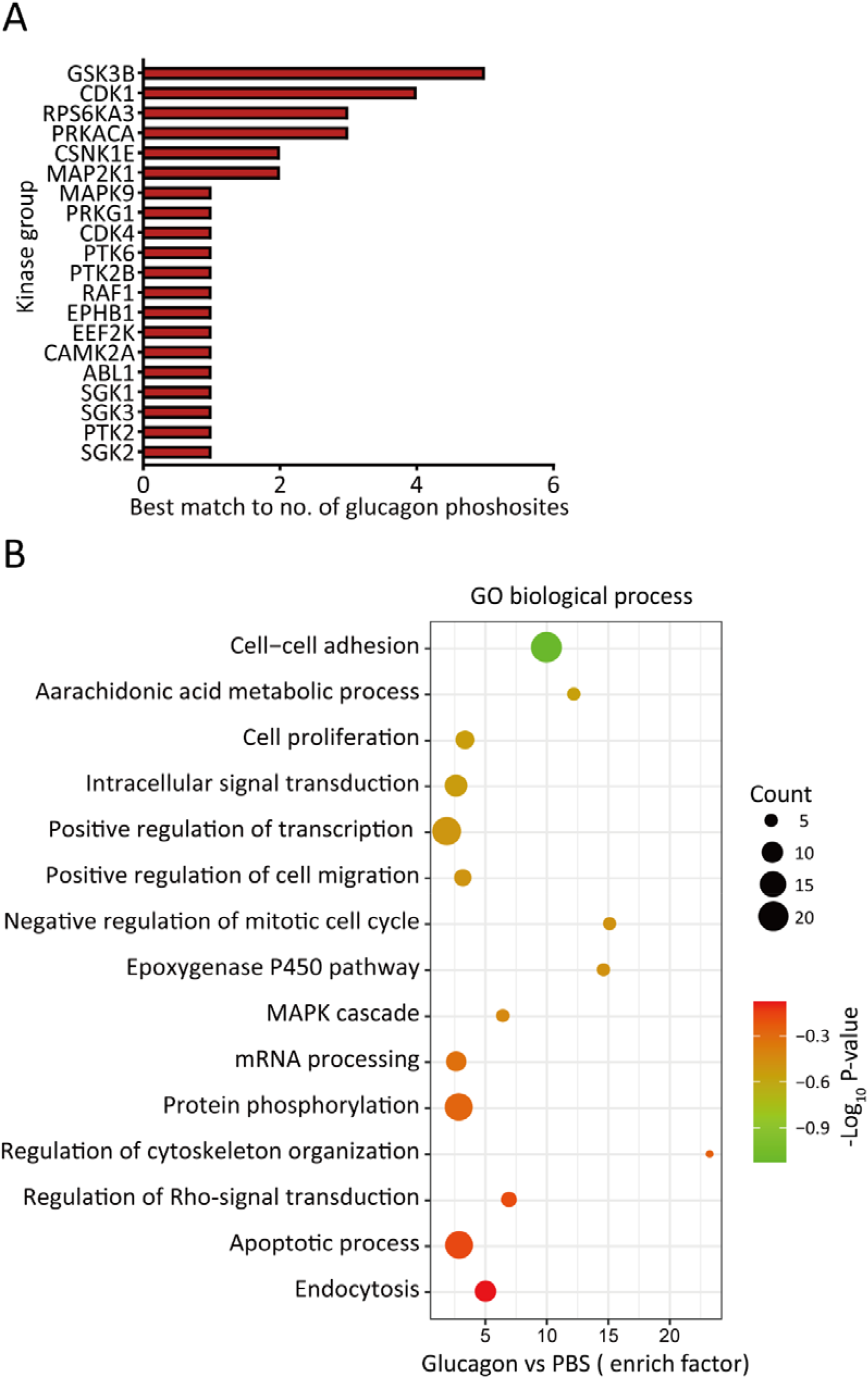
GP73 induces drastic remodeling of the PKA hub (related to Figure 5) (A) Distribution of matching kinases according to the phosphoproteomics data from the glucagon-treated sample (P <0.05) using Kinase Enrichment Analysis 2 (KEA2). (B) GO enriched pathway analysis of significantly regulated phosphopeptides in PMHs treated with rmGP73 (P <0.05) using DAVID Bioinformatics Resources 6.8. The bar plot shows significantly dysregulated pathways, and Fisher’s exact test P values are shown on the x-axis.

**Fig. S5.**
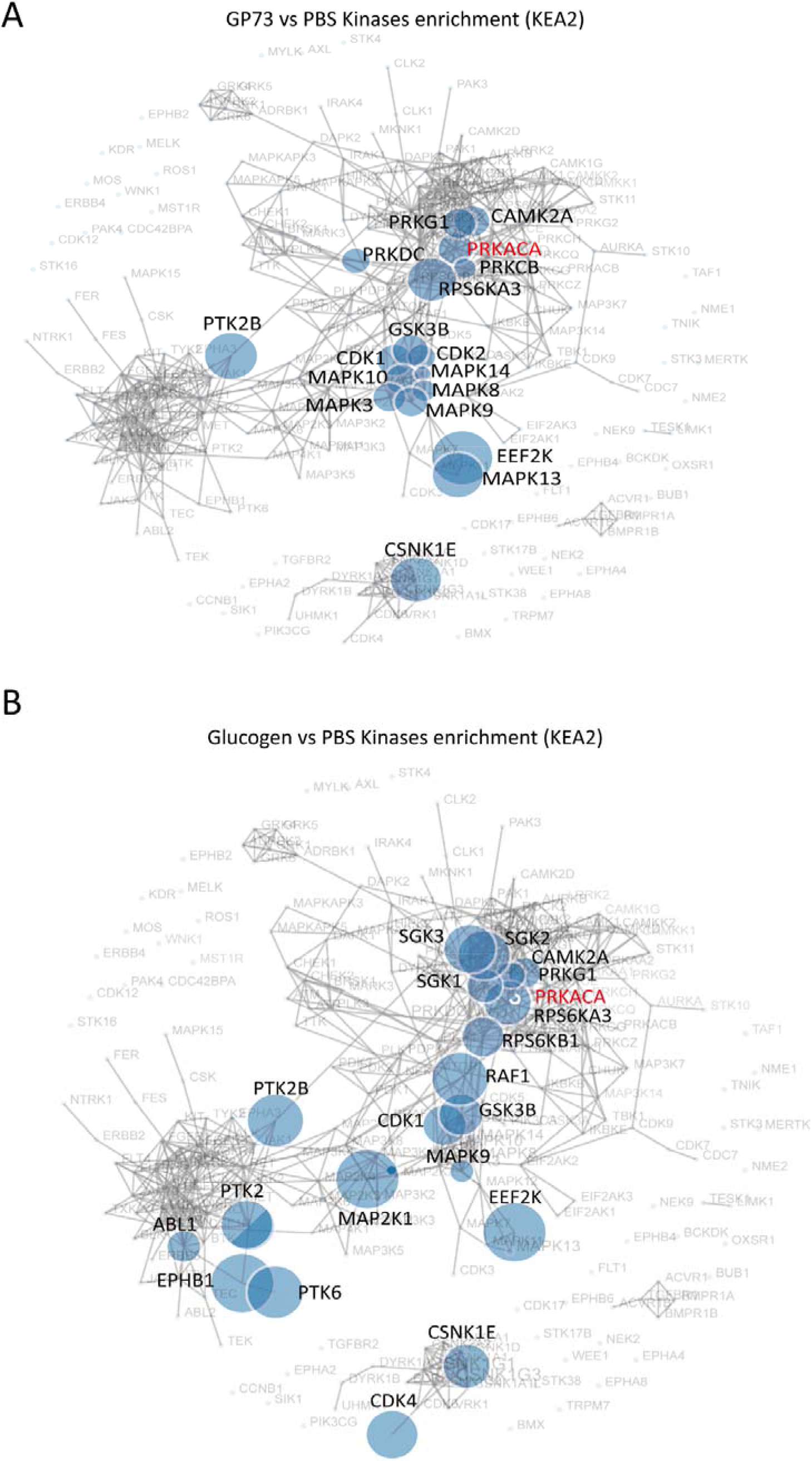
GP73 induces drastic remodeling of the PKA kinase hub (related to Figure 5) (A-B) The kinase activity network was constructed according to the phosphoproteomics data from the GP73-(A) or glucagon-(B) treated samples using Kinase Enrichment Analysis 2 (KEA2).

**Fig. S6.**
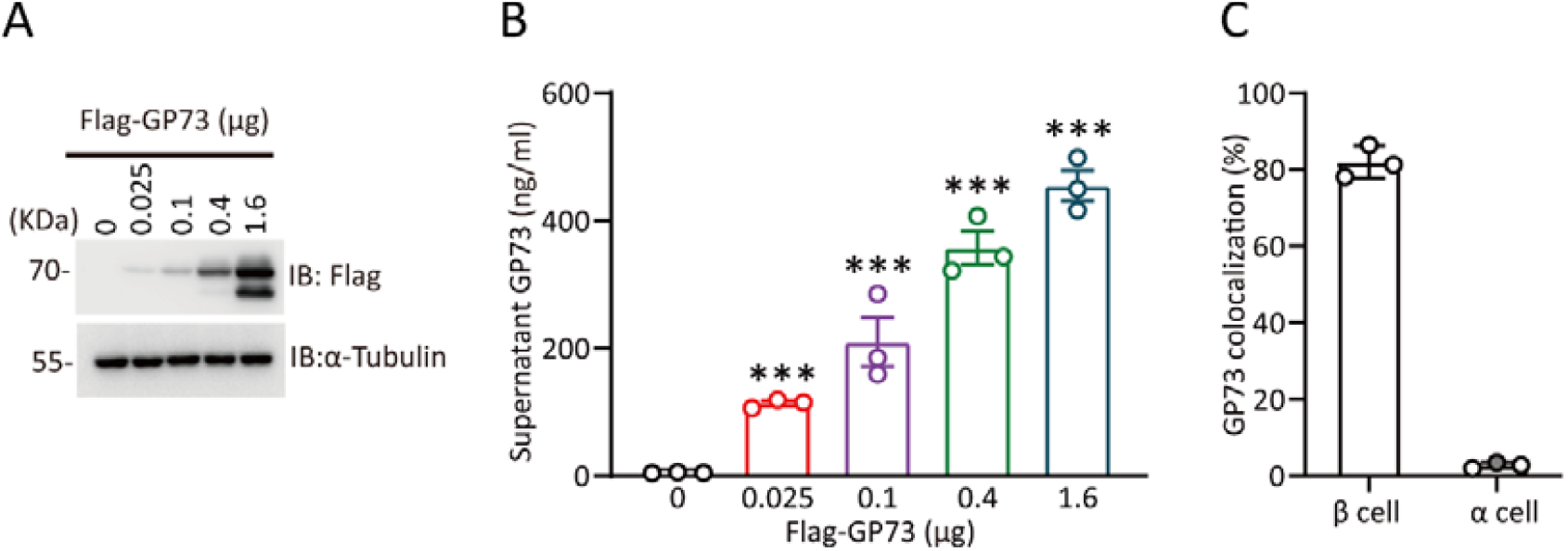
GP73 secretion is induced from multiple tissues upon fasting (related to Figure 6) (A-B) Immunoblotting of intracellular GP73 (A) and ELISA of supernatant GP73 levels (B) in HepG2 cells transfected with different concentrations of Flag-GP73. (C) Quantification of GP73 and insulin colocalization (β cells) and GP73 and glucagon colocalization (α cells) in Figure 6G.

**Fig. S7.**
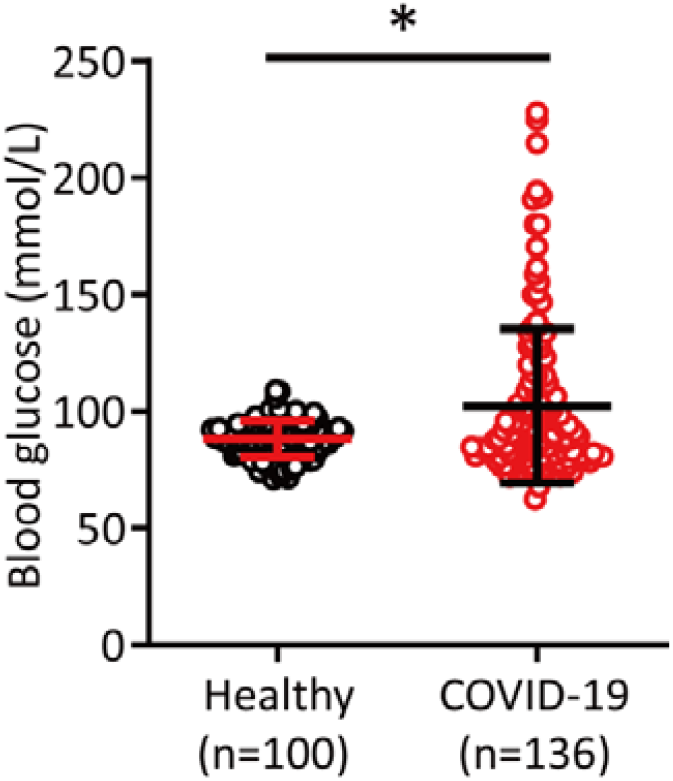
GP73 secretion is promoted under glucotoxicity conditions associated with SARS-CoV-2 infection and diabetes (related to Figure 7) Blood glucose levels in healthy controls (n=100) and COVID-19 patients. *, P < 0.05; Mann-Whitney U test.

**Fig. S8.**
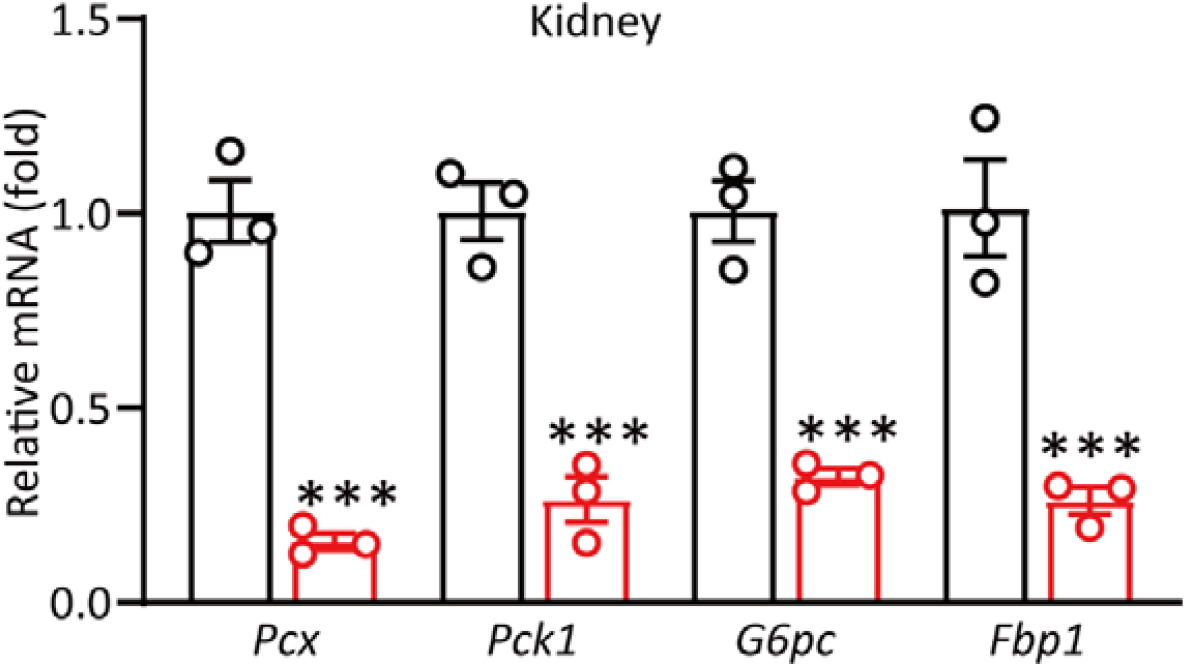
GP73 blockade reduces gluconeogenesis and lowers hyperglycemia associated with diabetes (related to Figure 9) QRT-PCR analysis of glucogenic gene expression in the kidneys of WT and GP73 KO adult mice fasted for 6 h (n=3 in each group). The data are the means ± SEM. ***, P < 0.001; one-way ANOVA followed by Bonferroni’s post hoc test.

## STAR Methods

### Lead Contact, Materials Availability and Data and Code Availability

Further information and requests for resources and reagents should be directed to and will be fulfilled by the Lead Contact Hui Zhong. All unique/stable reagents generated in this study are available from the Lead Contact with a completed Materials Transfer Agreement. The number of replicates carried out for each experiment is described in the figure/table legends.

### Reagents

Sodium L-lactate (71718), sodium pyruvate (792500), streptozotocin (STZ, S0103) and aprotinin (A6106) were purchased from Sigma-Aldrich (Missouri, USA). lnsulin (2018283062) was purchased from Novo Nordisk (Bagsværd, Denmark). Glucagon (HY-P0082) and FSK (HY-15371) were purchased from MedChemExpress (New Jersey, USA). Glucose (20171108) was purchased from Sinopharm Chemical Reagent (Beijing, China). A blood glucose meter (06656919032) and test strips (1072332990) were purchased from Roche (Basel, Switzerland). Sulfo-Cyanine7 NHS ester (Cy7, GY1058) was purchased from Goyoo Biotechnology (NanJing, China). Dulbecco’s modified Eagle medium (DMEM, high-glucose, D5796), protease inhibitor cocktail I (20-201) and a dipeptidyl peptidase-4 (DPP4) inhibitor (DPP4-010) were purchased from Millipore (Massachusetts, USA). Low-glucose DMEM (31600-500), glucose-free DMEM (90113-500) and L-alanine (A8210) were purchased from Solarbio (Beijing, China). Fetal bovine serum (FBS, A3160901) was purchased from Gibco (New York, USA). Lipofectamine™ 2000 (11668-027) was purchased from Invitrogen (Massachusetts, USA). Chow (HD1001) and a HFD (HD001) were purchased from BiotechHD (Beijing, China). cAMPS-Rp triethylammonium salt (151837-09-1) was purchased from Tocris Bioscience (Bristol, England). Collagenase IV (2091) was purchased from BioFroxx (Einhausen, Deutschland). PerfectStart™ Green qPCR SuperMix (AQ601) and TransScript® One-Step gDNA Removal and cDNA Synthesis SuperMix (AT311) were purchased from TransGen Biotech (Beijing, China). NucleoZOL (740404) was purchased from MACHEREY-NAGEL (MN, Düren, Deutschland). A mouse insulin ELISA kit (PI602) and BCA protein concentration determination kit (P0012) were purchased from Beyotime (Shanghai, China). A mouse glycated plasma protein kit (80420) was purchased from Crystal Chem (Washington, USA). Mouse aspartate transaminase (AST, 200218), alanine transaminase (ALT, 191230), triglyceride (TG, 200224) and cholesterol (CHO, 200224) biochemical test kits were purchased from Ruierda Biological Technology (Beijing, China). A cAMP assay kit (ab133051) and PKA Kinase Activity Kit (ab139435) were purchased from Abcam (Illinois, USA). An Amplex™ Red Glucose/Glucose Oxidase Assay Kit (A22189) was purchased from Invitrogen (Massachusetts, USA).

### Antibodies

Anti-α-tubulin (T6074, 1:5,000 dilution) and anti-Flag (A8592, 1:5,000 dilution) antibodies were purchased from Sigma-Aldrich (Missouri, USA). Anti-glucagon (ab92517, 1:2500 dilution), anti-insulin (ab6995, 1:200 dilution), anti-CREB-phospho S133 (ab32096, 1:1000 dilution), and anti-CREB (ab32515, 1:1000 dilution) antibodies were purchased from Abcam (Illinois, USA). An anti-GP73 antibody (F-12, sc-393372, 1:200 dilution) was purchased from Santa Cruz (Texas, USA). An anti-His antibody (KM8001, 1:1000 dilution) was purchased from Taihua Lekang Biotechnology (Beijing, China). Anti-phospho-Akt (Thr308, 13038, 1:1000 dilution), anti-Akt (9272, 1:1000 dilution), anti-phospho-PKA-C-α (Thr197, 5661, 1:1000 dilution), anti-phospho-PKA substrate (RRXpS/T, 9624, 1:1000 dilution), and anti-PKA-C-α (5842, 1:1000 dilution) antibodies were purchased from Cell Signaling Technology (Danvers, USA). Anti-rabbit HRP-IgG (ZB-2301, 1:5000 dilution) and anti-mouse HRP-IgG (ZB-2305, 1:5000 dilution) secondary antibodies were purchased from ZSGB-BIO (Beijing, China). An anti-GP73 monoclonal antibody (mAb) for the blocking experiment was custom made. Isotype-matched IgG (A7028) was purchased from Beyotime (Shanghai, China).

### Plasmids and cell culture

Mammalian expression vectors encoding Flag-tagged human, mouse and rat GP73 were constructed by inserting the corresponding PCR-amplified fragments into pcDNA3 (Invitrogen, Massachusetts, USA). The HepG2 (CRL-10741), Vero E6 (CRL-1568), HK-2 (CRL-2190), 293T (CRL-3216) and L6 (CRL-1658) cell lines were obtained from the American Type Culture Collection (ATCC, Rockville, MD, USA). The Huh-7 (0403) cell line was obtained from the Japanese Collection of Research Bioresources. All cell lines tested for mycoplasma contamination were incubated in DMEM at 37°C in a humidified atmosphere with 5% CO_2_. Lipofectamine™ 2000 was used for transfection following the manufacturer’s protocol.

To knock out human GP73 in Huh-7 cells, two small guide RNAs (sgRNAs) targeting GP73 were designed and inserted into the LentiCrispr v2 vector to construct transfer plasmids. 293T cells were transfected with pMD2. G, psPAX2 and the corresponding transfer plasmid to produce lentivirus. A total of 10^8^ Huh-7 cells were infected with lentivirus at a multiplicity of infection (MOI) of 2.0 and selected with 4 μg/mL puromycin for two weeks to ensure proper selection.

The following sgRNA sequences were used:

sgRNA-1: 5′-CACCGCACACACAGAGGTGCCACAA-3′

sgRNA-2: 5′-CACCGACCAGTTAAAGACCCTGCAG-3′

control-5′-CACCGCGCTTCCGCGGCCCGTTCAA-3′.

PMHs were isolated and purified using a modified two-step collagenase perfusion method. Cells were resuspended in low-glucose DMEM containing 5% FBS and seeded on 15-cm dishes at 80% confluence. Five hours later, the cells were washed and cultured in serum-free medium overnight. For gluconeogenesis-related assays, the medium was replaced with glucose- and phenol-free DMEM the next day in the presence of 10 mM pyruvate sodium and 10 mM sodium lactate, and the cells were treated with the indicated concentrations of rmGP73 or rrGP73, 200 mM cAMPS-Rp, or 2 μM glucagon. Then, 64 nM GP73 was used for 10 min for the cAMP and PKA assays and for 1 h for phosphoproteomics. The results were normalized to the protein content.

### Sample acquisition from COVID-19 patients

The Ethics Committee of Huoshenshan Hospital approved the study (HSSLL036). Given the urgency of the COVID-19 pandemic, the need for informed consent forms was waived by the ethics boards of the hospitals. Basic information and serum biochemical test results were collected from 136 COVID-19 patients at Huoshenshan Hospital (Wuhan, Hubei province, People’s Republic of China) from January 11 to March 11, 2020. Diagnosis was based on chest computed tomography (CT) manifestations and/or reverse transcription-polymerase chain reaction (RT-PCR) according to the criteria of the New Coronavirus Pneumonia Prevention and Control Program (5th edition) published by the National Health Commission of China.

According to these criteria, COVID-19 patients were classified into mild, moderate, and severe COVID-19 subgroups. Data were excluded if the subject was younger than 18 years or older than 75 years, had incomplete medical records, acute lethal organ injury (e.g., acute myocardial infarction, acute coronary syndrome, acute pulmonary embolism, or acute stroke), or decompensated or end-stage chronic organ dysfunction (e.g., decompensated cirrhosis, decompensated chronic renal insufficiency, or severe congestive heart failure), were pregnant or had malignancy (Table 1). Human blood samples were collected from the COVID-19 patients analyzed in this study before intervention. Fifty patients were classified as having mild COVID-19, 65 patients had moderate COVID-19, and 21 patients had severe COVID-19 (Table 2).

### Sample acquisition from healthy and diabetic patients

Human blood samples from the healthy and diabetic patients used in this study were obtained from individuals admitted to the Third Medical Center of the Chinese PLA General Hospital. The detailed characteristics of the recruited subjects are described in Table 3. All individuals in this study provided a signed statement of consent. The Committee for Ethics in Human Studies from the Third Medical Center of the Chinese PLA General Hospital approved this study (KY2021-009).

### Infection with authentic SARS-CoV-2

The SARS-CoV-2 strain (2019-nCoV BetaCoV/Beijing/AMMS01/2020) used in the present study was isolated from the lung lavage fluid of an infected patient and preserved at the State Key Laboratory of Pathogen and Biosecurity at Beijing Institute of Microbiology and Epidemiology. SARS-CoV-2 infections were performed in the BSL-3 Laboratory of the Beijing Institute of Microbiology and Epidemiology. The infectious virus titer was determined as plaque-forming units in Vero E6 cells and was used to calculate the MOI. Cells were infected with SARS-CoV-2 at the indicated MOIs for the indicated times in glucose-free medium for the production assay. The results were normalized to the protein content. Cells were infected with SARS-CoV-2 at the indicated MOIs for the indicated time, and the supernatant was harvested for the GP73 assay and viral load assay. Viral RNA was extracted from the supernatants using the QIAamp Viral RNA Mini Kit (52906, Qiagen) according to the manufacturer’s instructions. Viral RNA was analyzed using qRT-PCR and a One-Step PrimeScript RT-PCR Kit (RR064B, Takara) using SARS-CoV-2-specific primers in an Applied Biosystems 7500 Real-time PCR System. The following sequence of the SARS-CoV-2 probe was used:

SARS-CoV-2 open reading frame 1b (ORF1b):

Forward: 5′-CCCTGTGGGTTTTACACTTAA-3′

Reverse: 5′-ACGATTGTGCATCAGCTGA-3′.

Probe: 5′-FAM-CCGTCTGCGGTATGTGGAAAGGTTATGG-BHQ 1-3′.

SARS-CoV-2 nucleocapsid (N):

Forward: 5′-GGGGAACTTCTCCTGCTAGAAT-3′

Reverse: 5′-CAGACATTTTGCTCTCAAGCTG-3′.

Probe: 5′-FAM-TTGCTGCTGCTTGACAGATT-BHQ 1-3′.

The number of copies per microliter (μl) was determined using a synthetic RNA fragment to amplify the target region.

### Recombinant GP73 protein purification

Human, mouse, and rat GP73 cDNAs, each with a six-amino-acid His tag on the N-terminus, were cloned into the pCDNA3.1 vector. Ni-NTA His-Bind column-bound His-GP73 protein from 293T cells transfected with the above plasmids was further purified using size-exclusion columns and polymyxin B-based endotoxin-depletion columns after extensive washing. The final His-GP73 proteins used in all recombinant protein experiments were >90% pure (endotoxin<=2 EU/ml) and stored at -80°C.

### Animals, intervention and monitoring

Male C57BL/6N WT mice (8 to 10 weeks old) were purchased from SPF Biotechnology (Beijing, China). GP73 KO mice (T20200316-18[D25]) were generated by Southern Model Biotechnology (Shanghai, China). Male C57 BLKS/J db/db and BKS control mice (8 weeks, 36-40 g) were purchased from GemPharma Tech Co. Ltd. (Jiangsu, China). All mice were group-housed conventionally on a 12-h light/dark cycle for 3 days before any experiments. All animal experiments were performed at the AMMS Animal Center (Beijing, China) and were approved by the Institutional Animal Care and Use Committee. For single injections, mice were injected i.v. with 0.1 mg/kg recombinant His-tagged GP73, and plasma was collected at the indicated times via tail bleeding for insulin and glucose level measurements. The ITT, GTT, PTT, and ATT were performed using standard procedures. A 0.75 U/kg insulin bolus was used for the ITT, a 1.5 g/kg glucose bolus was used for the GTT, a 1.5 g/kg pyruvate bolus was used for the PTT, and a 0.6 g/kg alanine bolus was used for the ATT. For immunological sequestration experiments, mice were injected i.v. with 15 mg/kg custom-made anti-GP73 mouse mAb or an equivalent dose of IgG (30 mg/kg).

To establish an HFD-induced STZ model, mice were maintained on a regular chow diet or fed an HFD for 1 month beginning at 4 weeks of age. STZ (40 mg/kg) in citric acid buffer (0.1 mol/L, pH 4.2) was administered to male C57BL/6N mice via intraperitoneal injection, and the same dose of STZ was injected 24 h later. After STZ injection, the mice were fed an HFD for another month. Fasting blood glucose and random blood glucose levels were measured weekly. The mice were confirmed to have diabetes if their fasting blood glucose levels were over 199.8 mg/dL or their random blood glucose levels were over 300.6 mg/dL. For all experiments, mice were randomly assigned to different groups to ensure an unbiased distribution.

### Glucose measurement

All blood samples were collected from the tail, and glucose levels were measured using the glucose oxidase method and an automated blood glucose reader (ACCU-CHEK, Roche). For the measurement of fasting blood glucose levels, normal mice were fasted for 6 or 12 h, as indicated. Random blood glucose levels were measured at 9:00 a.m. If the glucose level was greater than 630 mg/dL (upper detection limit of the glucometer), a value of 630 mg/dL was recorded. Blood glucose levels were determined.

### Assays of plasma hormone levels

Blood samples for hormone detection were collected from the tail or orbital vein. A DPP4 inhibitor (1:100 dilution), aprotinin (1:100 dilution) and protease inhibitor cocktail I (50000 KIU/mL, 1:100 dilution) were added to each blood sample. Plasma insulin levels were measured using ELISA.

### Immunofluorescence staining

Tissues were fixed with 10% (v/v) neutral-buffered formalin at 4°C overnight and embedded in paraffin, and 5-μm-thick sections were prepared. For immunofluorescence, the sections were heated in an autoclave in citrate buffer (12 mmol/L, pH 6.0), preincubated in permeabilization/blocking buffer (0.1 mmol/L PBS, pH 7.3, 0.5% Triton) and blocked for 30 min with 10% (v/v) goat serum (Zhongshan Biotechnology, Beijing, China). The sections were subsequently incubated with primary antibodies at 4°C overnight and secondary antibodies for 1 h at room temperature, washed and stained with DAPI (1 μg/mL). Images were captured under a confocal fluorescence microscope (Zeiss LSM710, Carl Zeiss Microscopy GmbH, Jena, Germany) or an automatic digital slide scanner (Pannoramic MIDI, 3D HISTECH, Budapest, Hungary).

### Phosphoproteomics

PMHs were suspended in low-glucose DMEM containing 5% FBS and seeded in 15-cm dishes at 80% confluence. The cells were washed and cultured in serum-free medium overnight 5 h later. The medium was replaced with glucose- and phenol-free DMEM supplemented with 10 mM pyruvate sodium and 10 mM sodium lactate the next day, and the cells were incubated for 1 h with PBS, rmGP73 (64 nM), or glucagon (2 μM). For cell lysate collection, the cells were washed twice with cold PBS and scraped with cold RIPA lysis buffer supplemented with protease and phosphatase inhibitors. Phosphoproteomics was performed by Oebiotech Company. Briefly, the samples were subjected to enzyme digestion and iTRAQ labeling, and the phosphopeptides were enriched and analyzed using LC-MS/MS. The raw data of this study have been deposited in the IProX database with the following accession number: PXD025381. For kinase enrichment, the Literature Based Kinase-Substrate Library with Phosphosites on the Kinase Enrichment Analysis 2 (KEA2) website was searched. The network was represented using Cytoscape ver 3.6.2 (https://cytoscape.org/). Protein-protein interactions were retrieved from STRING App (v1.51) (https://string-db.org/). Only interactions with high confidence (interaction score >0.7) from databases and experiences were kept. For KEGG and GO enrichment analysis, DAVID Bioinformatics Resources 6.8 (https://david.ncifcrf.gov/home.jsp) was used. For specific kinase substrate motif analysis, MoMo Modification Motifs 5.3.3 (http://meme-suite.org/tools/momo) was used.

### In vivo imaging system (IVIS)

GP73 was labeled with Cy7 via the addition of the dye according to the manufacturer’s instructions at a pH of 8.0 and incubation of the mixture for 4 h on ice. The labeled GP73 was returned to a pH of 7.0, and the free dye was removed via overnight dialysis in PBS. The labeled GP73 was added to a Sephadex G50 size exclusion column equilibrated with PBS. Fractions of 500 µL were collected, the protein concentration was analyzed using a standard BCA protein assay kit, and fluorescence corresponding to excitation/emission of 745/800 nm was assessed. After i.v. injections with free Cy7 and GP73-Cy7, mice were scanned using an IVIS (PerkinElmer) at the indicated time points to assess fluorescence. After whole-body imaging, the mice were sacrificed, and the major organs were imaged to assess fluorescence under the same settings as the in vivo imaging. The data were analyzed and exported using built-in Living Image Software (Version 4.5.5, PerkinElmer).

### Quantitative real-time PCR (qRT-PCR)

Total mRNA was extracted from cells or various mouse tissues using NucleoZOL. cDNA was prepared from total mRNA using TransScript® One-Step gDNA Removal and cDNA Synthesis SuperMix, and the relative levels of individual mRNAs were calculated after normalization to the GAPDH mRNA level in the corresponding sample as previously described. The primer sequences are presented in Table 4.

**Table 4.**
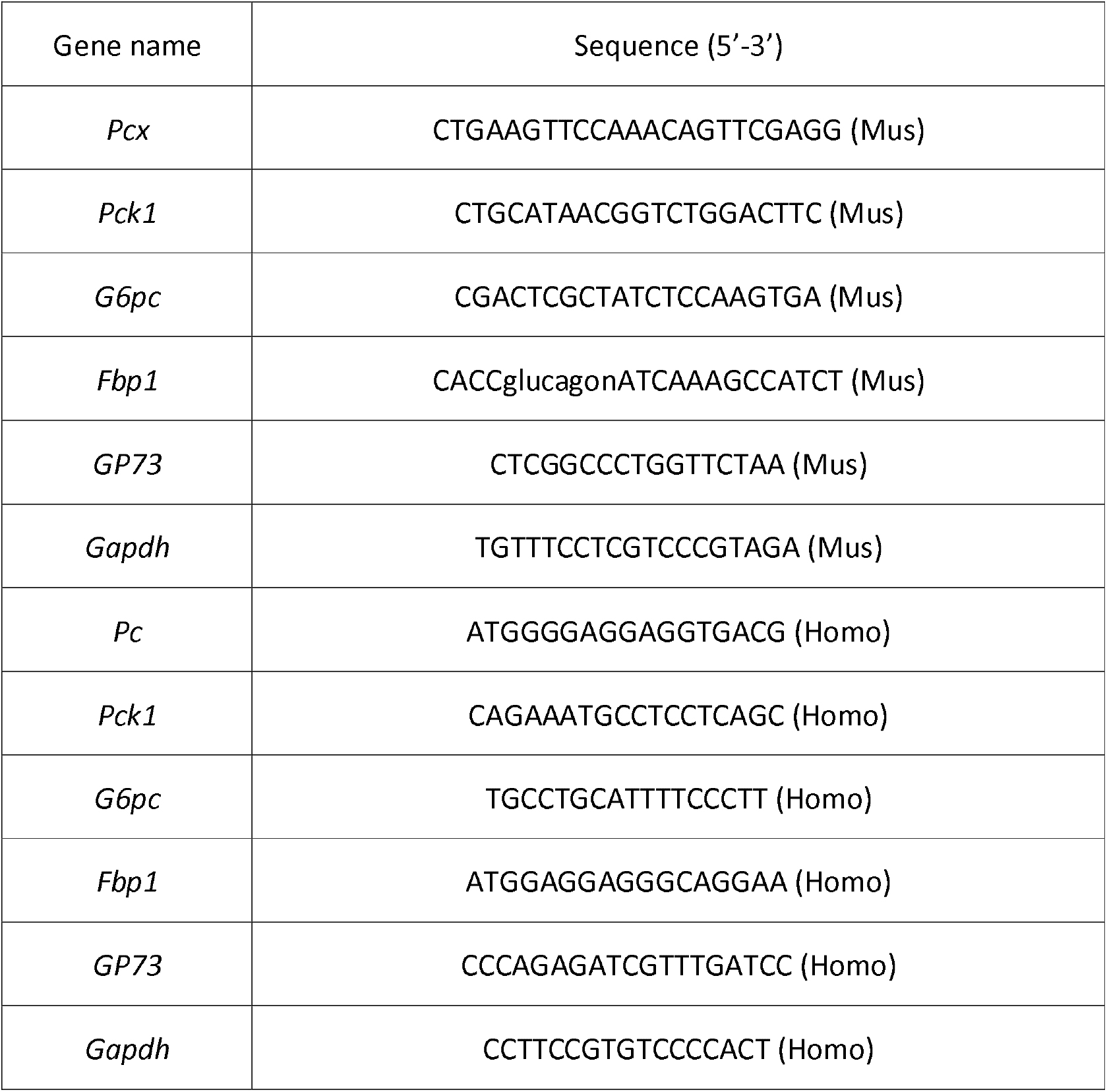
List of PCR primers used.

### Sandwich ELISA and Western blot analysis

For endogenous GP73 sandwich ELISA, two custom-made rat monoclonal anti-GP73 antibodies were used as the capture antibody and the detection antibody. Briefly, the plate was coated with an unlabeled capture antibody, and serially diluted standards and samples were added to the plate. After three washes, HRP-linked detection antibody was added to generate a colorimetric signal at 450 nm. For His-tagged GP73 sandwich ELISA, the same procedure was used, except a goat anti-His polyclonal antibody (Abcam) was used as the detection antibody. Increasing amounts of recombinant His-tagged GP73 were used to generate a standard curve for both assays.

For immunoblotting, cells were lysed in NP40 cell lysis buffer with fresh protease inhibitors. Whole-cell lysates were separated using SDS-PAGE after centrifugation and transferred to PVDF membranes for immunoblot analyses using the indicated primary antibodies.

### Statistical analysis

The present study used GraphPad Prism 8.0 for statistical calculations and data plotting. Differences between two independent samples were evaluated using two-tailed Student’s t-tests or the Mann-Whitney test, as appropriate. Differences between multiple samples were analyzed using one-way ANOVA or two-way ANOVA followed by Bonferroni’s post hoc test, as appropriate. Correlations were analyzed using Spearman’s non-parametric test. All tests were two-tailed unless otherwise indicated. We considered a P value ≤ 0.05 as statistically significant. Significance values were set as follows: ns (not significant), P > 0.05; *, P < 0.05; **, P < 0.01; and ***, P < 0.001.

